# Pathways through which intermittent preventive treatment for malaria in pregnancy influences child growth faltering: a mediation analysis

**DOI:** 10.1101/2024.06.09.24308656

**Authors:** Yanwei Tong, Kalani Ratnasiri, Suhi Hanif, Anna T. Nguyen, Michelle E. Roh, Grant Dorsey, Abel Kakuru, Prasanna Jagannathan, Jade Benjamin-Chung

## Abstract

**Background:** Intermittent preventive treatment for malaria in pregnancy (IPTp) can improve birth outcomes, but whether it confers benefits to postnatal growth is unclear. We investigated the effect of IPTp on infant growth in Uganda and its pathways of effects using causal mediation analyses.

**Methods:** We analyzed data from 633 infants born to mothers enrolled in a randomized trial of monthly IPTp with dihydroartemisinin-piperaquine (DP) vs sulfadoxine-pyrimethamine (SP) (NCT 02793622). Weight and length were measured from 0-12 months of age. Using generalized linear models, we estimated effects of DP vs. SP on gravidity-stratified mean length-for-age (LAZ) and weight-for-length Z-scores (WLZ). We investigated mediation by placental malaria, gestational weight change, maternal anemia, maternal inflammation-related proteins, preterm birth, birth length, and birth weight. Mediation models adjusted for infant sex, gravidity, gestational age at enrollment, maternal age, maternal parasitemia at enrollment, education, and wealth.

**Findings:** SP increased LAZ by 0.18-0.28 Z from birth through age 4 months compared to DP, while DP increased WLZ by 0.11-0.28 Z from 2-8 months compared to SP among infants of multigravidae. We did not observe these differences among primigravida. Mediators of SP included increased birth weight and length and maternal stem cell factor at delivery. Mediators of DP included placental malaria and birth length, maternal IL-18, CDCP1, and CD6 at delivery.

**Interpretation:** In high malaria transmission settings, different IPTp regimens influenced infant growth among multigravidae through distinct pathways in the period of exclusive breastfeeding, when few other interventions are available.

**Funding:** Stanford Center for Innovation and Global Health, Eunice Kennedy Shriver National Institute of Child Health and Human Development, Bill & Melinda Gates Foundation

**Research in context:** *Evidence before this study:* Intermittent Preventive Treatment in Pregnancy (IPTp) with sulfadoxine-pyrimethamine (SP) is recommended by the WHO for regions with moderate-to-high malaria transmission. While SP is effective in reducing neonatal mortality and low birth weight, its efficacy has diminished in some areas of sub-Saharan Africa due to widespread parasite resistance to SP. Although IPTp with dihydroartemisinin-piperaquine (IPTp-DP) has demonstrated superior efficacy in reducing malaria in pregnancy, its impact on birth outcomes has not significantly surpassed that of SP. The ultimate goal of IPTp extends beyond enhancing birth outcomes to include benefits during infancy and later stages. Yet, the effects of SP vs. DP in relation to infant growth post-birth and the underlying mechanisms remain unknown. Prior studies also found that different IPTp regimens worked through different pathways, with DP influencing birth outcomes by reducing placental malaria and SP influencing them through non-malarial pathways such as maternal weight gain. Here, we re-analyzed data from of a randomized trial in Uganda to explore the impacts of these two IPTp regimens on infant growth and to understand potential mechanisms underlying its impacts on infant growth.

*Added value of this study:* This study quantified how IPTp with SP compared to DP influenced infants’ growth trajectories, both ponderal and linear, during the first year of life. We found that SP improved linear growth of infants up to age 4 months compared to DP, and DP improved ponderal growth of infants from 2-8 months compared to SP among babies who were born to multigravidae. In addition, we identified birth size, placental malaria, and certain markers of maternal inflammation measured at delivery using the Olink Target 96 inflammation panel as pathways through which IPTp influenced infant growth. Our approach provides new insights into effects of IPTp beyond birth and the mechanisms by which IPTp impacts infant growth.

*Implications of all the available evidence:* Our study provides evidence that different IPTp regimens can influence infant postnatal growth through distinct pathways. Our findings highlight the potential of combined SP and DP IPTp regimens and bolster the evidence base for continued delivery of IPTp to improve maternal and child health outcomes, particularly in malaria-endemic regions.

## Introduction

Child growth faltering is associated with a large burden of disease, including increased risk of death and infections in childhood and lower productivity in adulthood.^1–4^ Children in low-and middle income countries often experience growth faltering before age 6 months, when complementary feeding and most child nutrition interventions are initiated.^2,5,6^ In malaria-endemic settings, prenatal malaria infection may be an important contributor to growth failure because of its effects on inflammation, anemia, and intrauterine growth restriction,^7–9^ which are linked to low birth weight, premature birth, stillbirth, and fetal death.^10–12^

In moderate-to-high *Plasmodium falciparum* (*Pf*) malaria transmission settings, the World Health Organization recommends intermittent preventive treatment of malaria in pregnancy (IPTp) with sulfadoxine-pyrimethamine (IPTp-SP).^13^ However, in eastern and Southern Africa, the antimalarial efficacy of SP has waned due to increasing parasite resistance to SP.^14^ Three trials in areas of high SP resistance of Kenya and Uganda found that IPTp with dihydroartemisinin-piperaquine (IPTp-DP) significantly reduced the risk of malaria during pregnancy, but were not associated with better birth outcomes relative to IPTp-SP.^15–17^ Ultimately, the purpose of IPTp is not only to improve outcomes at birth but also in infancy and beyond. Yet no prior studies have assessed effects of IPTp on child growth.

SP and DP likely influence child growth through distinct pathways given DP’s higher efficacy against malaria^15–17^ and SP’s antibiotic properties.^18^ Prior studies reported that effects of IPTp on birth weight were mediated by placental malaria for DP;^19^ given that maternal malaria infection is associated with impaired child height and weight gain in infancy, DP’s benefits may extend into childhood.^20–23^ Another study found that SP’s effect on birth weight was mediated by gestational weight gain,^24^ and maternal anthropometry is strongly associated with child growth.^2^ Maternal inflammation may be another important pathway: sequestration of *Pf* parasites in the placenta can result in inflammation, dysregulated development, and impaired nutrient transport in the placenta, which can negatively impact fetal development^25^ and child growth.^26–30^

Using data from a randomized trial of Ugandan mother-infant dyads, we assessed whether IPTp-DP improved infant growth from birth to 12 months compared to IPTp-SP. In addition, we investigated whether maternal inflammation, anemia, preterm birth, gestational weight gain, and birth weight and length mediated the effects of IPTp on child growth from birth to 12 months.

## Methods

### Study population

This is a secondary analysis of data collected in a double-blind randomized phase III trial in Busia district, a high malaria transmission area of southeastern Uganda (NCT02793622).

Between September 6, 2016, and May 29, 2017, the study enrolled 782 HIV-uninfected women, at least 16 years of age, with a viable pregnancy between 12 and 20 weeks of gestation confirmed by ultrasound. The study followed their live births for 12 months from April 1, 2017 to October 31, 2018. Eligible women with a history of serious adverse events to SP or DP, early or active labor, chronic medical conditions or active medical problems requiring inpatient evaluation, or previous antimalarial therapy during the pregnancy were not enrolled in the study. Women were randomized at a 1:1 ratio to receive monthly IPTp with SP or monthly DP starting at 16 or 20 weeks of gestation. Both pharmacists administering medications and study participants were masked. We excluded multiple births (e.g., twins), spontaneous abortions, and stillbirths. The final analysis dataset included data from 633 children in the birth cohort.

### Follow-up measurements

Women were scheduled for routine visits every 4 weeks. During each visit, blood was collected to detect the presence of malaria parasites by microscopy or quantitative PCR (qPCR). Women also underwent routine laboratory testing every 8 weeks. Children born to the study participants were scheduled for visits to the clinic at 1, 4, 6, and 8 weeks of age and then every 4 weeks until they reached 52 weeks of age. Women were encouraged to visit the study clinic for all medical care for themselves or their children; the clinic was open daily, and participants received a refund for transportation costs.

Study participants, both mothers and their children, had a standardized history and physical exam taken during clinic visits which included temperature, pulse, and blood pressure measurements. Those who were febrile (tympanic temperature > 38.0 °C) or reported a fever in the past 24 hours and had a positive blood smear were treated for malaria with artemether-lumefantrine for uncomplicated malaria and intravenous artesunate for complicated malaria.

### Intervention

DP was given as 3 tablets taken once a day for 3 consecutive days every month (40 mg dihydroartemisinin and 320 mg piperaquine; Duo-Cotecxin, Holley-Cotec, Beijing). SP was given as a single dose consisting of 3 tablets every month (500 mg of sulfadoxine and 25 mg of pyrimethamine; Kamsidar, Kampala Pharmaceutical Industries, Uganda). To ensure participant blinding, participants in the DP arm received SP placebos, and participants in the SP arm received DP placebos each month. Study staff directly observed administration of the first dose of each intervention. Subsequent doses were self-administered at home, and adherence for those doses was assessed by self-report.

### Mediators

Mediators included gestational weight change, prenatal anemia, placental malaria, preterm birth, low birth weight, and maternal inflammation. Since pre-pregnancy weight was not available, instead of measuring gestational weight gain, we calculated the change in gestational weight from 20 to 36 weeks gestation. We used weight at 36 weeks instead of delivery to increase consistency between women since the majority of women delivered after 36 weeks.

Staff measured maternal hemoglobin at 28 weeks gestation and classified mothers as anemic if blood hemoglobin was < 11 g/dL. Placental malaria was assessed at delivery using histopathological assessment of placenta tissue or testing for parasitemia in the placental blood by loop-mediated isothermal amplification. In the analysis, we considered women who had placental malaria under either method to be positive. We classified low birth weight as birth weight ≤2500g and preterm birth as deliveries prior to 37 weeks gestation.

Using maternal plasma samples collected at delivery in a random, arm-stratified subsample of 264 mothers, we measured inflammation-related proteins using the Olink Target 96 inflammation panel. This high-multiplex immunoassay panel identifies 92 proteins associated with immune response. Inflammation biomarker data were log2 transformed and normalized to a Normalized Protein eXpression (NPX) relative quantification unit such that a one-unit change in NPX is equivalent to a two-fold increase in protein concentration. Twenty-five proteins were excluded from analysis because they failed to quantify in >50% of samples or the NPX value was below its limit of detection in >50% of samples (Table S1). We used principal components analysis and established pathway analysis and term enrichment databases (Blood Transcriptional Modules, Gene Ontology, KEGG) to reduce the dimensionality and identify clusters of proteins. However, we did not observe meaningful clusters, so statistical analyses used individual proteins. We corrected p-values for Olink analyses using Benjamini-Hochberg correction (false discovery rate p-value < 0.05).^31^

### Outcomes

Study staff measured length and weight in infants each month from birth to 12 months. We calculated Z-scores using the World Health Organization (WHO) Child Growth Reference Standard.^32^ We calculated mean length-for-age (LAZ), weight-for-length (WLZ), and weight-for-age (WAZ) Z-scores in quarterly intervals. In a sensitivity analysis, we corrected Z-scores for gestational age: for infants with gestational age < 37 weeks, we calculated age as the postnatal age subtracted by difference between gestational age at birth and 280 days.

We defined stunting as LAZ < 2 standard deviations (SD) below the median of the WHO standard and wasting as WLZ < 2 SDs below the median. For binary outcomes, we calculated incidence in quarterly intervals as the number of children who became stunted or wasted during the interval divided by the number who were not previously stunted or wasted at the start of the interval. We assumed children were at risk of each outcome at birth or their first measurement.

### Statistical analysis

The analysis plan was pre-specified at https://osf.io/f8wy4/. See deviations in Supplement 1. To understand the mechanisms through which IPTp affects child growth faltering, we used causal mediation analyses (Figure S1).^33–35^ We estimated total effects as the effect of IPTp on child growth faltering through all pathways, including through measured and unmeasured mediators. The total effect can be decomposed into the direct effect (i.e., “natural direct effect”) and the mediated effect (i.e., “natural indirect effect”).^36^ The direct effect measures the effect of IPTp on child growth if we disabled pathways through mediators, while the mediated effect measures the effect of IPTp on child growth that operates through mediators.

We estimated total effects of IPTp DP vs. SP using generalized linear models with a Gaussian family and identity link for continuous outcomes (Z-scores) and generalized linear models with a Poisson family and log link for dichotomous outcomes (stunting and wasting).^37^ We fit unadjusted models because characteristics were balanced at baseline between study groups.^17^

We fit models to estimate effects of IPTp (DP vs. SP) on mediators and associations between mediators and each outcome using generalized linear models as defined above. We report crude intervention-mediator effects since the intervention was randomized. For mediation analyses, mediator and outcome models adjusted for mediator-outcome confounders by choosing nodes sufficient to block backdoor pathways in Figure S1. Potential confounders included maternal gravidity, infant sex, gestational age at enrollment, maternal age, maternal parasitaemia status at enrollment, education, and household wealth. We estimated mediation parameters and obtained 95% confidence intervals using a quasi-Bayesian Monte Carlo approach with 1,000 simulations.^38^ We compared outcome models with and without intervention-mediator interaction terms; because results were similar, we report estimates from models without interaction terms. We performed a complete case analysis. The percentage of live births with anthropometric measurements was 98% at birth, 89% at 6 months, and 78% at 12 months; follow-up was similar between arms.

Identification assumptions of causal mediation analyses include no unmeasured confounding of the intervention-outcome relationship, mediator-outcome relationship, or intervention-mediator relationship; temporal ordering intervention, mediators, and outcomes; and no mediator-outcome confounder that is itself affected by the intervention.^34^ The first assumption is met because intervention was randomized. We adjusted for confounders in intervention-outcome and mediator-outcome models to minimize confounding. Intervention, mediators, and outcomes were temporally sequenced.

### Ethics

This study was approved by the Institutional Review Board at Stanford University (#40857); the original trial was approved by ethics committees of Makerere University School of Biomedical Sciences (Kampala, Uganda, approval number SBS-342), the Uganda National Council for Science and Technology (Kampala, Uganda; HS 2052).

### Role of funding source

The funders of the study had no role in study design, data collection, data analysis, data interpretation, or writing of the report. The corresponding author had full access to all the data in the study and had final responsibility for the decision to submit for publication.

## Results

### Characteristics of the study population

The study initially enrolled 782 women. This analysis excluded 149 individuals due to study withdrawal, spontaneous abortions, stillbirth, non-singleton birth, and missing placental malaria measurements. The analysis sample included 633 singleton mother-infant dyads (Figure S2), 152 (24%) of which were primigravidae. Mothers’ mean age was 24 years. The average gestational weight change between week 20 and week 36 was 3.4 kg. We documented 296 cases of anemia and 34 (5%) preterm deliveries. The prevalence of placental malaria at delivery was 80% among primigravidae and 33% among multigravidae.

### Child growth faltering

At birth, median LAZ was −1.0 (IQR=1.05) and median WLZ was 0.55 (IQR=1.63); mean LAZ was −0.99 (95% CI −1.07, −0.91) and mean WLZ was 0.49 (95% CI 0.40, 0.59) (Figure 1a-b). The proportion of children experiencing incident stunting onset was 18% at birth and 23% from age 1 day to 3 months, and <10% at subsequent ages (Figure 1c). At each age, the incidence of severe stunting was 6% or lower, with higher incidence before age 3 months. Wasting incidence was 13% from 1 day to 3 months and was <4% at all other ages (Figure 1d). Severe wasting incidence was <3% at all ages.

**Figure 1.**
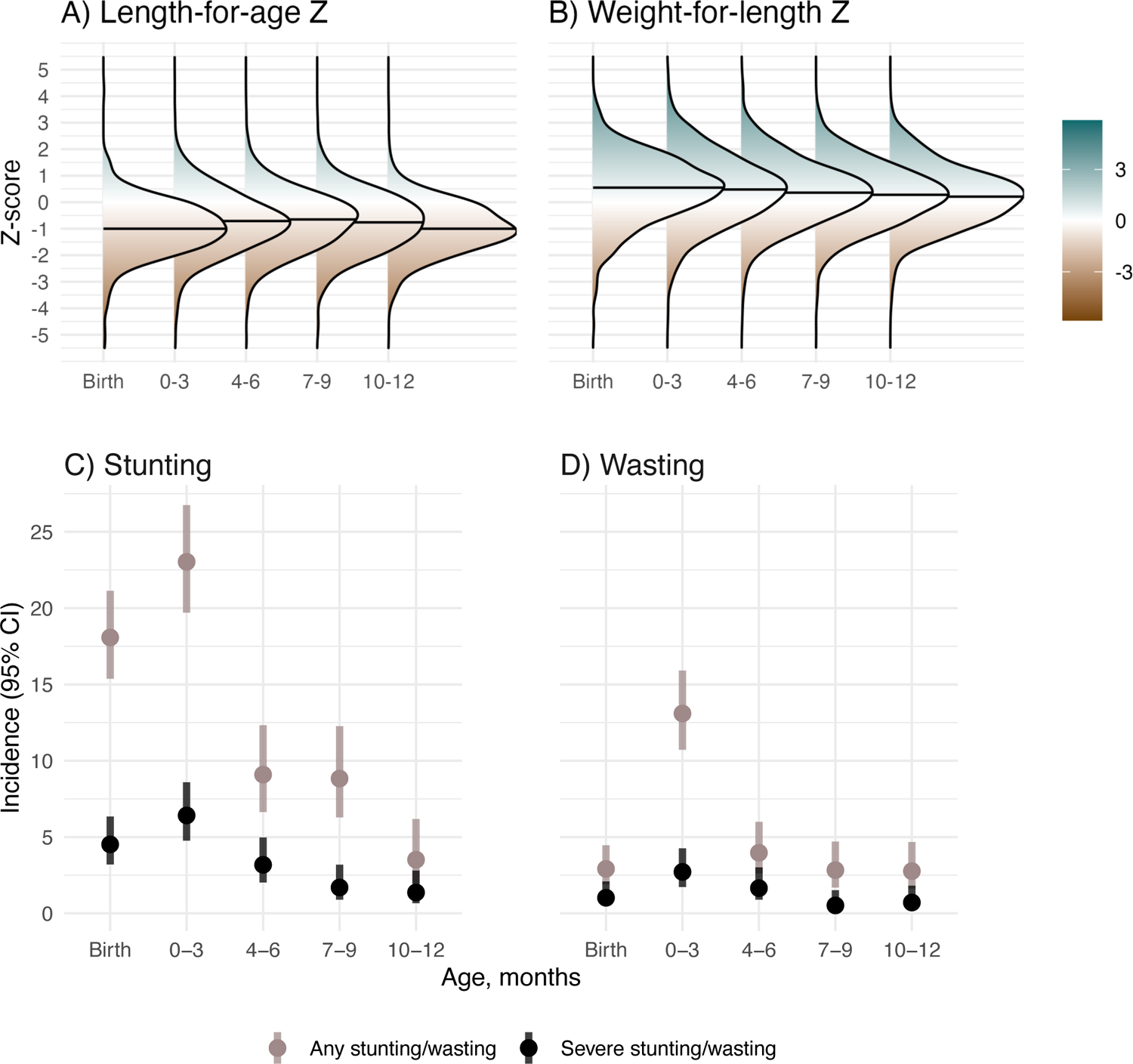
Child growth from birth through 12 months. 0-3 includes children aged 1 day to 3 months, 4-6 includes children aged >3 to 6 months, 7-9 includes children aged >6 to 9 months, and 10-12 includes children aged >9 to 12 months. In A) and B) horizontal lines in each density plot indicate the median Z-score for each age category.

### Effects of IPTp on child growth

Among primigravidae, there were minimal differences in LAZ and WLZ by age between study arms (Figure 2). Among multigravidae, SP increased mean LAZ by 0.19-0.27 Z from birth to 4 months compared to DP, with mean LAZ around −0.55 in the SP arm and −0.75 in the DP arm at these ages. Compared to SP, DP increased mean WLZ from 2-8 months by 0.11-0.28 Z. Gestational age correction increased LAZ by approximately 0.2 Z and WLZ by approximately 0.5 Z and did not change patterns by arm (Figure S3). The incidence of stunting and wasting by age was similar between study arms (Table S2).

**Figure 2.**
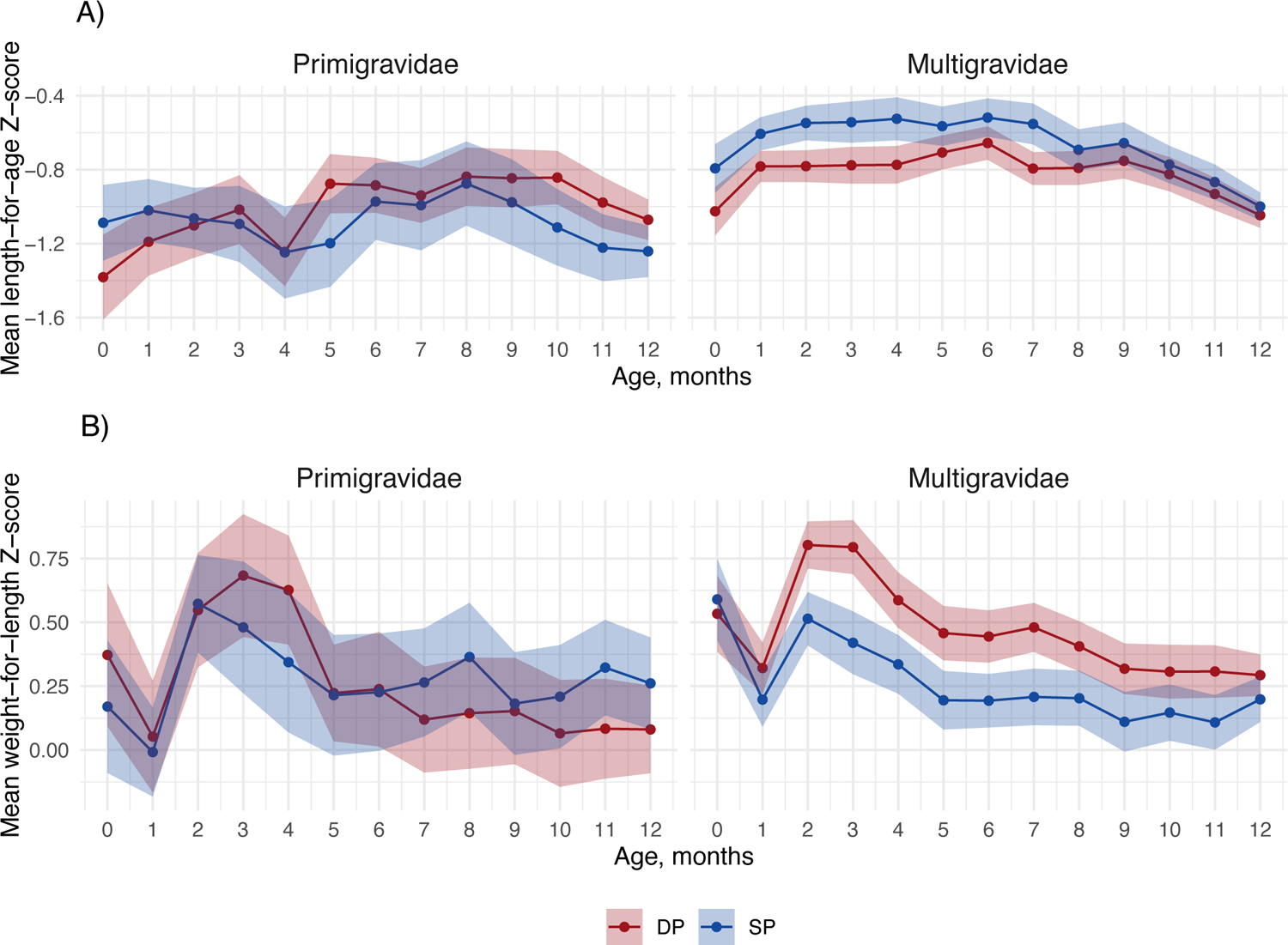
Total effect of IPTp DP vs. SP on mean child growth Z-scores by child age and gravidity Includes 633 children measured from birth to twelve months. The shaded region represents the 95% confidence interval, colored by IPTp group.

### Effects of IPTp on mediators

SP improved mean birth length by 0.47 cm (95% CI 0.11, 0.82) and mean birth weight by 60 g (95% CI 0, 130) compared to DP (Figure 3). For birth length, the effect was larger among primigravidae, but for birth weight, the effect was only present among multigravidae. Placental malaria was lower for DP compared to SP (incidence ratio = 0.45, 95% CI 0.35, 0.58), and the effect was stronger among multigravidae.

**Figure 3.**
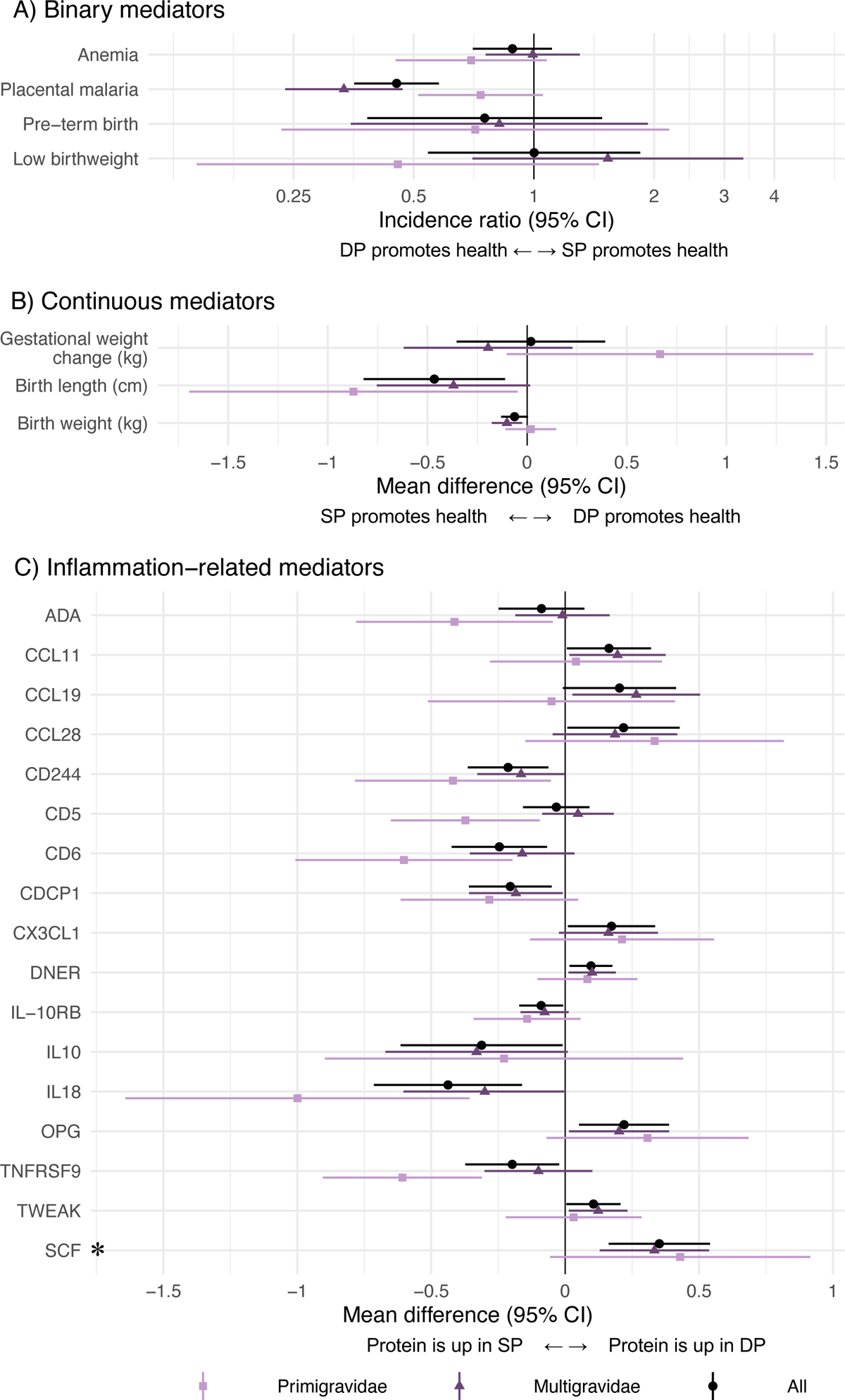
Effect of IPTp-DP vs. IPTp-SP on potential mediators Unadjusted incidence ratios and mean differences between IPTp-DP and IPTp-SP. The asterisk indicates a statistically significant effect in Olink inflammation analyses among all gravidae after Benjamini-Hochberg correction (false discovery rate p-value < 0.05). C) only includes Olink results with statistically significant differences between arms; results for all Olink proteins are shown in Figure S4.

We identified 17 biomarkers that were either upregulated in DP relative to SP or vice versa. DP increased certain inflammation-related proteins at delivery, including Fractalkine (CX3CL1), Delta and Notch-like epidermal growth factor-related receptor (DNER), Osteoprotegerin (OPG), and SCF relative to SP. SP increased CD244, T cell surface glycoprotein CD6 isoform (CD6), CUB domain-containing protein 1 (CDCP1), interleukin-18 (IL-18), and tumor necrosis factor receptor superfamily member 9 (TNFRSF9) compared to DP. The largest effects were on OPG, IL-18, and SCF. Compared to SP, DP increased SCF by 0.35 NPX (95% CI 0.16, 0.54), increased OPG by 0.24 NPX (95% CI 0.08, 0.41), and decreased IL-18 by 0.48 NPX (95% CI 0.20, 0.76). These are equivalent to 70%, 48%, and −96% percent changes, respectively. After applying the Benjamini-Hochberg correction, only stem cell factor (SCF) was upregulated in DP (Figure 3, Figure S4). We did not observe differences in anemia, pre-term birth, low birth weight, gestational weight change, or other inflammation-related proteins between arms.

### Association between mediators and child growth

Mediators strongly associated with LAZ and WLZ included pre-term birth, birth weight, and birth length (Figure S5). Placental malaria was associated with 0.30 (95% CI 0.03, 0.57) lower mean LAZ at birth among multigravidae, but there was no association at other ages or among primigravidae. Preterm birth was associated with increased stunting incidence at birth and increased stunting and wasting incidence from 1 day to 3 months (Table S3). Birth length was associated with lower stunting incidence from birth through 9 months and higher wasting incidence at birth. Birth weight was associated with lower stunting and wasting incidence at certain ages. Multiple inflammation-related proteins were associated with LAZ, WLZ, stunting, and wasting (Figure S6-S7). After multiple testing correction, only two protein associations with WLZ at certain ages were statistically significant (Figure S6).

### Mediators of the effect of IPTp on linear growth

Mediators of the effect of SP compared to DP on LAZ included birth weight and birth length and inflammation-related proteins SCF and DNER. Birth weight was a mediator from birth through 3 months, and birth length was a mediator at all ages (Figure 4a, Figure 5). Reductions in SCF mediated SP’s effects on LAZ at birth (Benjamini-Hochberg (BH) p=0.072), and reductions in DNER mediated effects at 6-9 months (BH p=0.504). Compared to SP, DP increased LAZ from 1 day-3 months by increasing OPG (BH p=0.396). The effect of IPTp on stunting at birth was mediated by pre-term birth, low birth weight, and SCF (BH p=0.543) (Figure S8). For linear growth outcomes, there was no evidence of mediation by placental malaria, maternal anemia, preterm birth, gestational weight change, or other inflammation-related proteins (Table S4, Figures S9, S10, S11).

**Figure 4.**
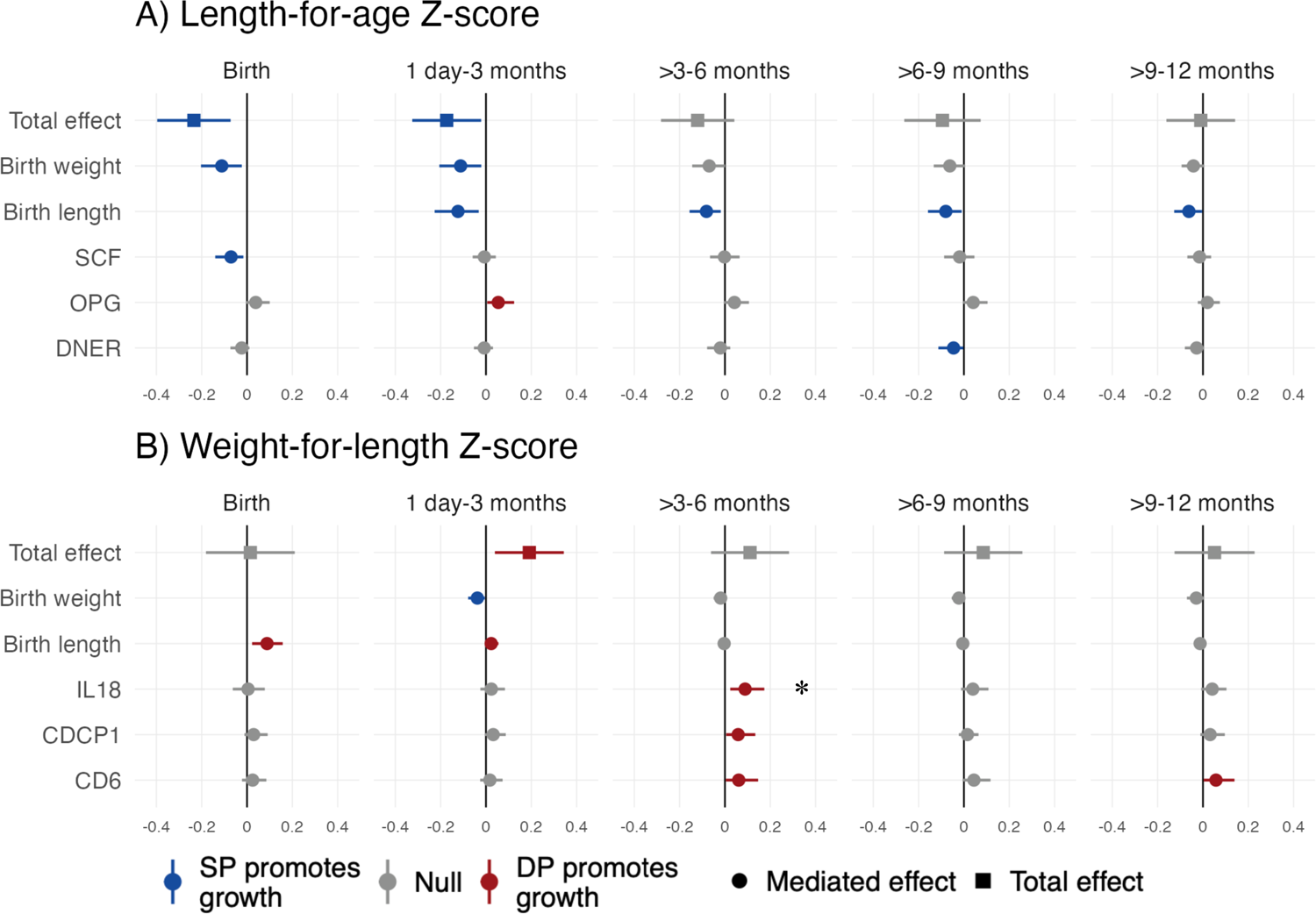
Total effects and mediated effects on child growth Z-scores The total effects compare mean Z-scores between IPTp-DP and IPTp-SP using unadjusted models. The mediated effects were adjusted by infant sex, maternal age, maternal baseline parasitemia, gestational age at enrollment, maternal education, household wealth, and gravidity. The reference group was SP. Includes all gravidae. *Statistically significant among all gravidae after Benjamini-Hochberg correction (false discovery rate p-value < 0.05).

**Figure 5.**
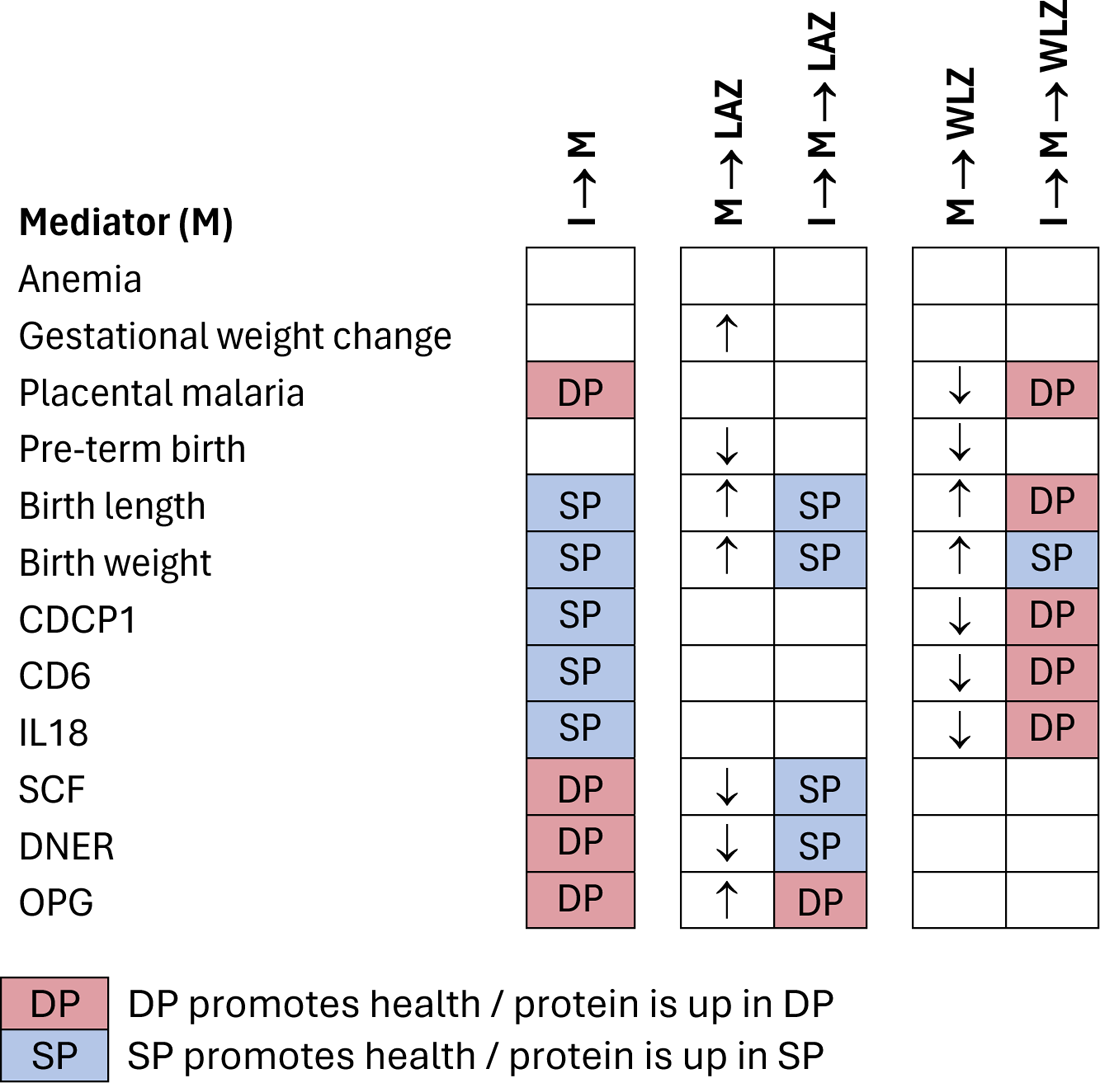
Summary of results for each mediating pathway I: Intervention (IPTp DP vs. SP); LAZ: length-for-age Z; WLZ: weight-for-length Z I ® M indicate results of intervention-mediator models. M ® LAZ indicate results of models of the association between each mediator and LAZ (and analogously for WLZ). I ® M® LAZ indicate results of mediation models for LAZ (and analogously for WLZ). Cells with DP indicate that IPTp-DP improved the outcome compared to IPT-SP or that DP increased inflammation-related proteins relative to SP. We considered improvements to be reductions in anemia, placental malaria, pre-term birth, and inflammation-related proteins and increases in gestational weight change and birth length or weight. Arrows indicate whether the mediator was associated with higher(τ) or lower (1) mean Z-scores.

### Mediators of the effect of IPTp on ponderal growth

Mediators of the effect of DP compared to SP on WLZ included birth size up to 3 months and certain inflammation-related proteins later in infancy. Relative to DP, SP increased WLZ from birth through 3 months by increasing birth weight (Figure 4b, Figure 5). DP’s effect on WLZ from birth to 3 months was mediated by birth length; this reflects lower birth lengths and greater weight gain in DP vs. SP. The effect of DP on increased WLZ at birth was also mediated by reduced placenta malaria relative to SP, however, the confidence interval included the null (Table S4). Relative to SP, DP improved WLZ by decreasing IL-18, CDCP1, and CD6 from 4-6 months; CD6 was also a mediator from 9-12 months. After multiple testing correction, the only BH p-value <0.05 was for IL-18 at 3-6 months. The effect of IPTp on wasting was mediated by low birth weight and birth weight up to age 3 months (Figure S8). There was no evidence of mediation of the effect of DP or SP on WLZ or wasting by placental malaria, maternal anemia, preterm birth, gestational weight change, or other inflammation-related proteins (Table S4, Figures S9, S12, S13).

## Discussion

In this secondary analysis of a randomized trial of Ugandan mother-infant dyads, we found that IPTp-SP improved linear growth from birth to age 4 months compared to DP, and IPTp-DP improved ponderal growth from ages 2 to 8 months compared to SP among infants born to multigravidae. IPTp regimens affected growth through distinct pathways: SP increased linear and ponderal growth via increased birth size. DP increased ponderal growth through reduced inflammation and reduced placental malaria, consistent with a prior analysis,^19^ though evidence was weak for the latter. SP’s effects on linear growth and DP’s effects on ponderal growth were mediated by reductions in different inflammation-related proteins, with the strongest evidence for mediation of DP’s effect on ponderal growth by reduced IL-18. We did not observe differences between IPTp regimens on child growth in primigravidae, possibly because the adverse consequences of placental malaria are more severe in this subgroup, which may have offset the ‘non-malarial’ benefits of SP.^19^

Overall, the effects on child growth among multigravidae were similar in size or larger than effect sizes of preventive nutritional interventions. IPTp-SP increased mean LAZ by up to 0.27 Z compared to DP, and DP increased mean WLZ by up to 0.28 Z compared to SP. These differences in Z are similar to effect sizes for infant multiple micronutrient supplements in the exclusive breastfeeding stage.^39^ Many other early life nutritional interventions, such as nutrition education and counseling,^40^ complementary feeding,^40^ maternal micronutrient supplements,^39^ and small quantity lipid-based nutrient supplements, have had smaller effects.^41^ Importantly, we observed benefits of IPTp to child growth during the exclusive breastfeeding stage, when growth faltering onset is high and few nutritional interventions are delivered.^5,6^

Certain inflammation-related proteins mediated the effects of IPTp on growth, with the strongest evidence for IL-18 as a mediator of DP and WLZ. IL-18 is a proinflammatory cytokine that is up-regulated during *Pf* malaria infection;^42,43^ it is present throughout pregnancy in the placenta^44^ and in the blood, where levels become elevated in labor.^45^ Increased levels of IL-18 are associated with early pregnancy loss, recurrent miscarriages and other pregnancy complications.^44–46^ Some studies suggest that maternal inflammation may impact infant growth outcomes.^27,47^ Taken together, our findings support the growing body of research pointing to maternal inflammation in pregnancy as a mediator of fetal and infant growth.

Potential mediating pathways not investigated in this study could also explain the observed differences in child growth between SP and DP. A separate analysis of this trial suggested that some of SP’s benefit may result from reduced febrile respiratory infections.^48^ Other studies point to SP’s effects on maternal nutrition: a trial found that SP increased maternal mid-upper arm circumference relative to DP,^49^ and an in vitro study found that SP improved nutrient absorption.^50^ Another trial identified gestational weight change as a mediator of the effect of IPTp-SP compared to DP.^24^ We did not observe mediation by gestational weight change, possibly because weight changes were relatively small in our study. In future studies, it would also be valuable to investigate whether SP’s antibiotic properties influence the vaginal or gut microbiome and whether SP modulates maternal immunity.^19^

Limitations include the relatively low incidence of stunting and wasting, particularly at older ages, which may have limited statistical power, particularly in primigravid women. Second, we only had data on inflammation-related proteins in a subsample of mothers at delivery, which could have confounded our findings due to the extensive inflammation that occurs during labor and delivery. Inflammation earlier in pregnancy may also be important. Third, it is possible that mediator-outcome models were subject to residual unmeasured confounding. Finally, we investigated mediators individually, which implicitly assumed that mediators operated independently; it is possible that some mediators operated jointly.

Our findings suggest that in high malaria transmission settings, different IPTp regimens influence infant growth through distinct pathways from birth through 6 months, when there is a dearth of other effective nutritional interventions. WHO IPTp policies have focused on low birth weight, but our findings suggest that different IPTp regimens could improve ponderal and linear growth beyond birth. In areas of high parasite resistance to SP, SP and DP likely improve infant growth through different mechanisms, warranting research on their combined use for enhanced public health impact compared to either regimen alone.

## Data and Code Availability

Data from the original trial is available at https://clinepidb.org/ce/app/workspace/analyses/DS_8786631aaf/new/variables/EUPATH_0000096/EUPATH_0015457. Replication scripts are available at https://github.com/YanweiTong-Iris/IPTp-BC3-mediation/tree/main

## Author contributions

JBC and PJ designed the study and obtained funding for this analysis. GD, AK, and PJ contributed to data collection in the original trial. JBC, SH, KR, and PJ developed the statistical analysis plan. JBC, YT, SH, and KR analyzed the data. YT, AN, KR, and JBC wrote the first draft of the manuscript. All authors reviewed and approved the final manuscript.

## Acknowledgements

This study was supported by the Stanford Center for Innovation in Global Health (270519). The original trial was supported by grants from the Eunice Kennedy Shriver National Institute of Child Health and Human Development (P01 HD059454) and the Bill & Melinda Gates Foundation. Research reported in this publication was supported in part by the National Institute of Allergy and Infectious Diseases of the National Institutes of Health under Award Numbers K01AI141616 (PI: Benjamin-Chung), U01 AI155325 (PI: Jagannathan), and the National Heart, Lung, And Blood Institute of the National Institutes of Health under award number T32HL151323 (Nguyen). MER is supported by the Eunice Kennedy Shriver National Institute of Child Health and Human Development Pathway to Independence Award (K99HD111572). The content is solely the responsibility of the authors and does not necessarily represent the official views of the National Institutes of Health. Jade Benjamin-Chung is a Chan Zuckerberg Biohub Investigator.

## SUPPLEMENT

**Table S1.** Inflammation-related proteins included in each analysis.

|  | Passed<br>LOD* check | Significant† intervention-mediator<br>results | Significant† mediator-outcome<br>results | Included in the mediation<br>analysis |
| --- | --- | --- | --- | --- |
| IL8 | √ |  |  |  |
| VEGFA | √ |  | √ |  |
| CD8A | √ |  | √ |  |
| MCP-3 |  |  |  |  |
| GDNF |  |  |  |  |
| CDCP1 | √ | √ | √ | √ |
| CD244 | √ | √ | √ | √ |
| IL7 | √ |  | √ |  |
| OPG | √ | √ | √ | √ |
| LAP TGF-beta-1 | √ |  | √ |  |
| uPA | √ |  | √ |  |
| IL6 | √ |  | √ |  |
| IL-17C | √ |  | √ |  |
| MCP-1 | √ |  | √ |  |
| IL-17A |  |  |  |  |
| CXCL11 | √ |  | √ |  |
| AXIN1 | √ |  | √ |  |
| TRAIL | √ |  | √ |  |
| IL-20RA |  |  |  |  |
| CXCL9 | √ |  |  |  |
| CST5 | √ |  | √ |  |
| IL-2RB |  |  |  |  |
| IL-1 alpha |  |  |  |  |
| OSM | √ |  | √ |  |
| IL2 |  |  |  |  |
| CXCL1 | √ |  | √ |  |
| TSLP |  |  |  |  |
| CCL4 | √ |  | √ |  |
| CD6 | √ | √ | √ | √ |
| SCF | √ | √ | √ | √ |
| IL18 | √ | √ | √ | √ |
| SLAMF1 |  |  |  |  |
| TGF-alpha | √ |  | √ |  |
| MCP-4 | √ |  | √ |  |
| CCL11 | √ | √ | √ | √ |
| TNFSF14 | √ |  | √ |  |
| FGF-23 |  |  |  |  |
| IL-10RA |  |  |  |  |
| FGF-5 |  |  |  |  |
| MMP-1 | √ |  |  |  |
| LIF-R | √ |  | √ |  |
| FGF-21 | √ |  | √ |  |
| CCL19 | √ | √ | √ | √ |
| IL-15RA |  |  |  |  |
| IL-10RB | √ | √ |  |  |
| IL-22 RA1 |  |  |  |  |
| IL-18R1 | √ |  | √ |  |
| PD-L1 | √ |  | √ |  |
| Beta-NGF |  |  |  |  |
| CXCL5 | √ |  | √ |  |
| TRANCE | √ |  | √ |  |
| HGF | √ |  | √ |  |
| IL-12B | √ |  | √ |  |
| IL-24 |  |  |  |  |
| IL13 |  |  |  |  |
| ARTN |  |  |  |  |
| MMP-10 | √ |  | √ |  |
| IL10 | √ | √ | √ | √ |
| TNF | √ |  | √ |  |
| CCL23 | √ |  | √ |  |
| CD5 | √ | √ | √ | √ |
| CCL3 | √ |  | √ |  |
| Flt3L | √ |  |  |  |
| CXCL6 | √ |  | √ |  |
| CXCL10 | √ |  | √ |  |
| 4E-BP1 | √ |  | √ |  |
| IL-20 |  |  |  |  |
| SIRT2 | √ |  | √ |  |
| CCL28 | √ | √ | √ | √ |
| DNER | √ | √ | √ | √ |
| EN-RAGE | √ |  | √ |  |
| CD40 | √ |  | √ |  |
| IL33 |  |  |  |  |
| IFN-gamma | √ |  | √ |  |
| FGF-19 | √ |  | √ |  |
| IL4 |  |  |  |  |
| LIF |  |  |  |  |
| NRTN |  |  |  |  |
| MCP-2 | √ |  | √ |  |
| CASP-8 | √ |  | √ |  |
| CCL25 | √ |  |  |  |
| CX3CL1 | √ | √ |  |  |
| TNFRSF9 | √ | √ | √ | √ |
| NT-3 |  |  |  |  |
| TWEAK | √ | √ | √ | √ |
| CCL20 | √ |  |  |  |
| ST1A1 | √ |  | √ |  |
| STAMPB | √ |  | √ |  |
| IL5 |  |  |  |  |
| ADA | √ | √ | √ | √ |
| TNFB | √ |  |  |  |
| CSF-1 | √ |  | √ |  |

\* LOD: limit of detection
† p-value not accounting for multiple testing < 0.05

### Supplement 1. List of deviations from pre-analysis plan

The analysis plan for this study was pre-specified at https://osf.io/f8wy4/. We note the following deviations from the plan:

1. We intended to employ principal components analysis and established pathway analysis and term enrichment databases, such as Blood Transcriptional Modules, Gene Ontology, and KEGG, to reduce the dimensionality and identify clusters of Olink inflammation-related biomarkers. However, these approaches did not yield discernible clusters. Therefore, we adhered to parametric regression, t-tests, volcano plots, and forest plots to investigate individual biomarkers. These approaches allowed us to systematically identify and track individual biomarkers demonstrating evidence of associations with the interventions and child growth outcomes for subsequent inclusion in our mediation models.
2. The hypothesized causal pathways of the study as depicted in Figure S1 were sequential. However, due to limitations in available R programming tools, we decided to focus on single-mediator models to examine each intervention-mediator-outcome combination separately.

---

**Figure S1.**
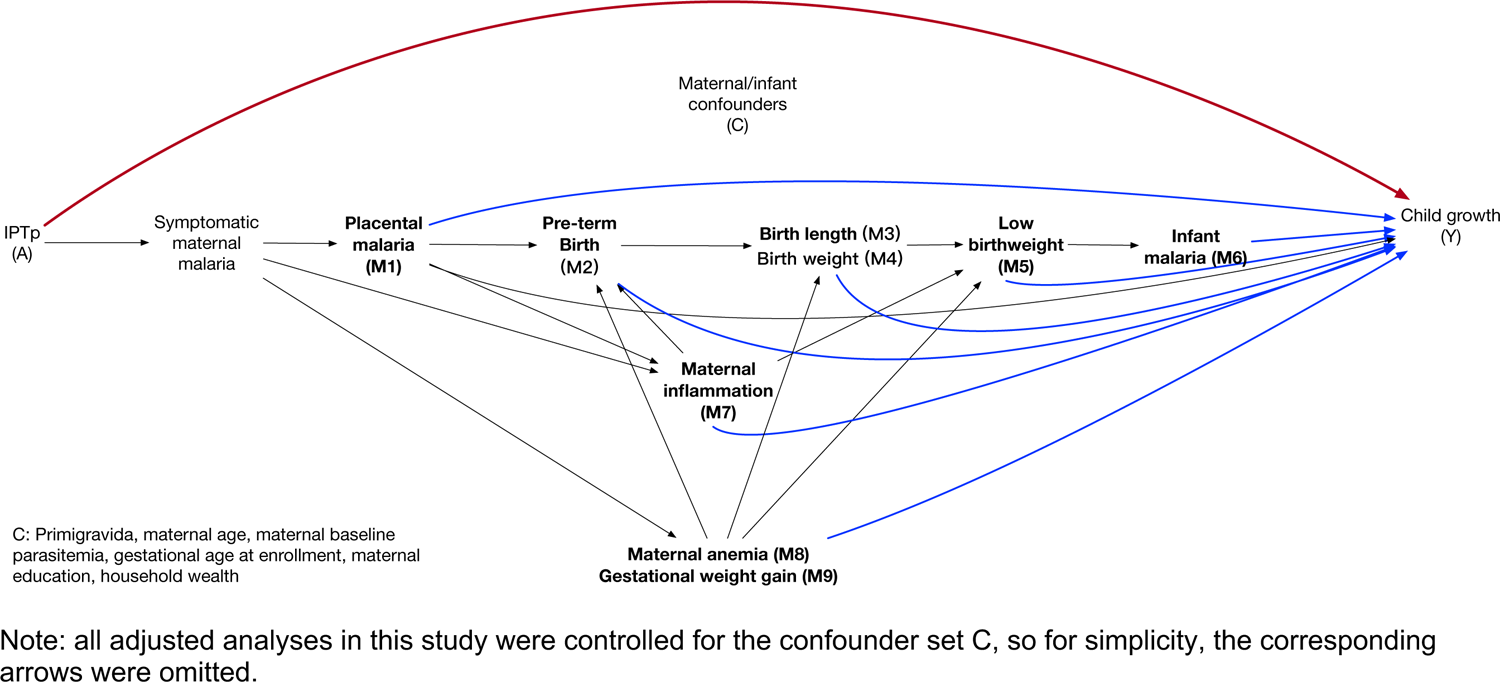
Directed acyclic graph

**Figure S2.**
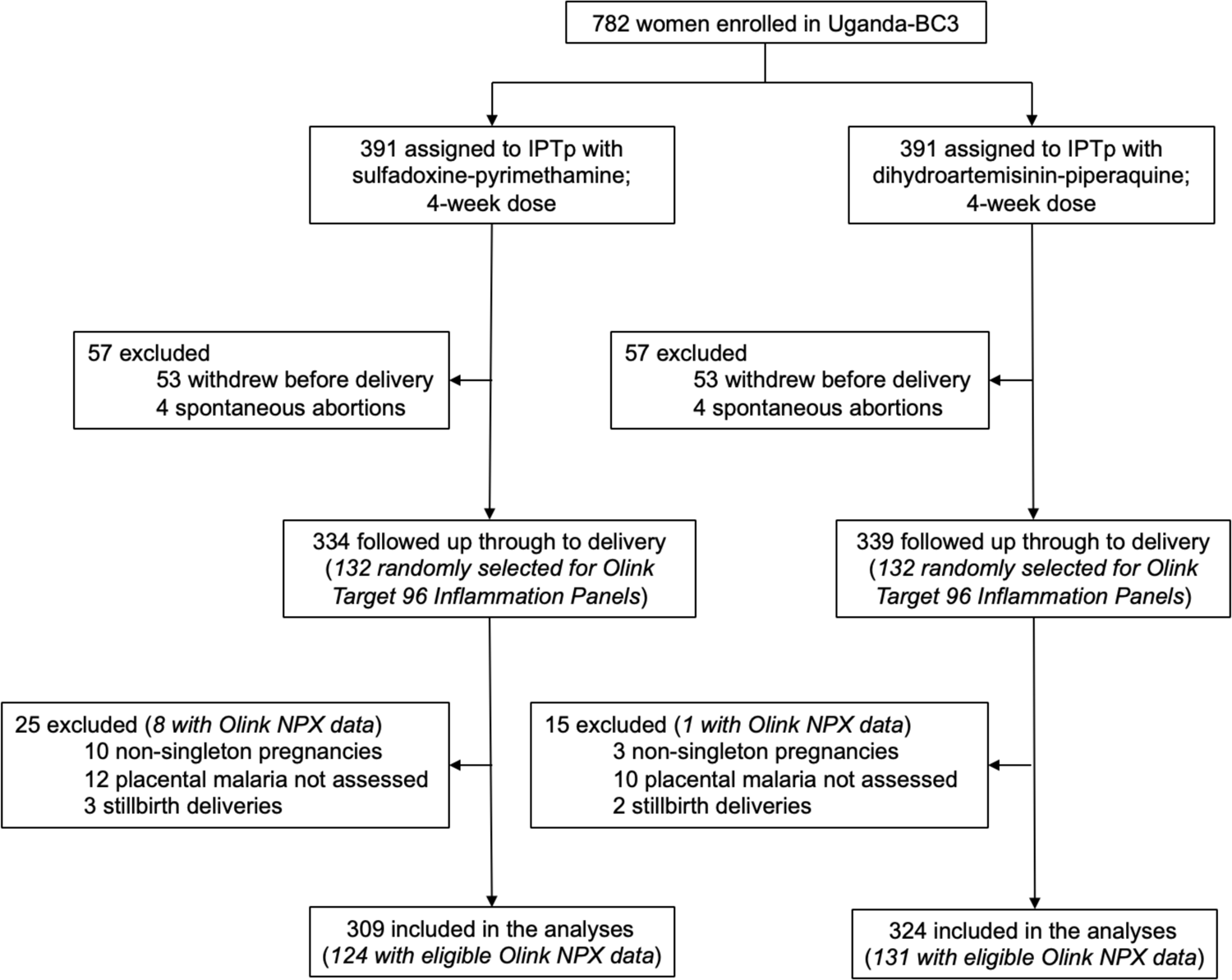
Flowchart

**Figure S3.**
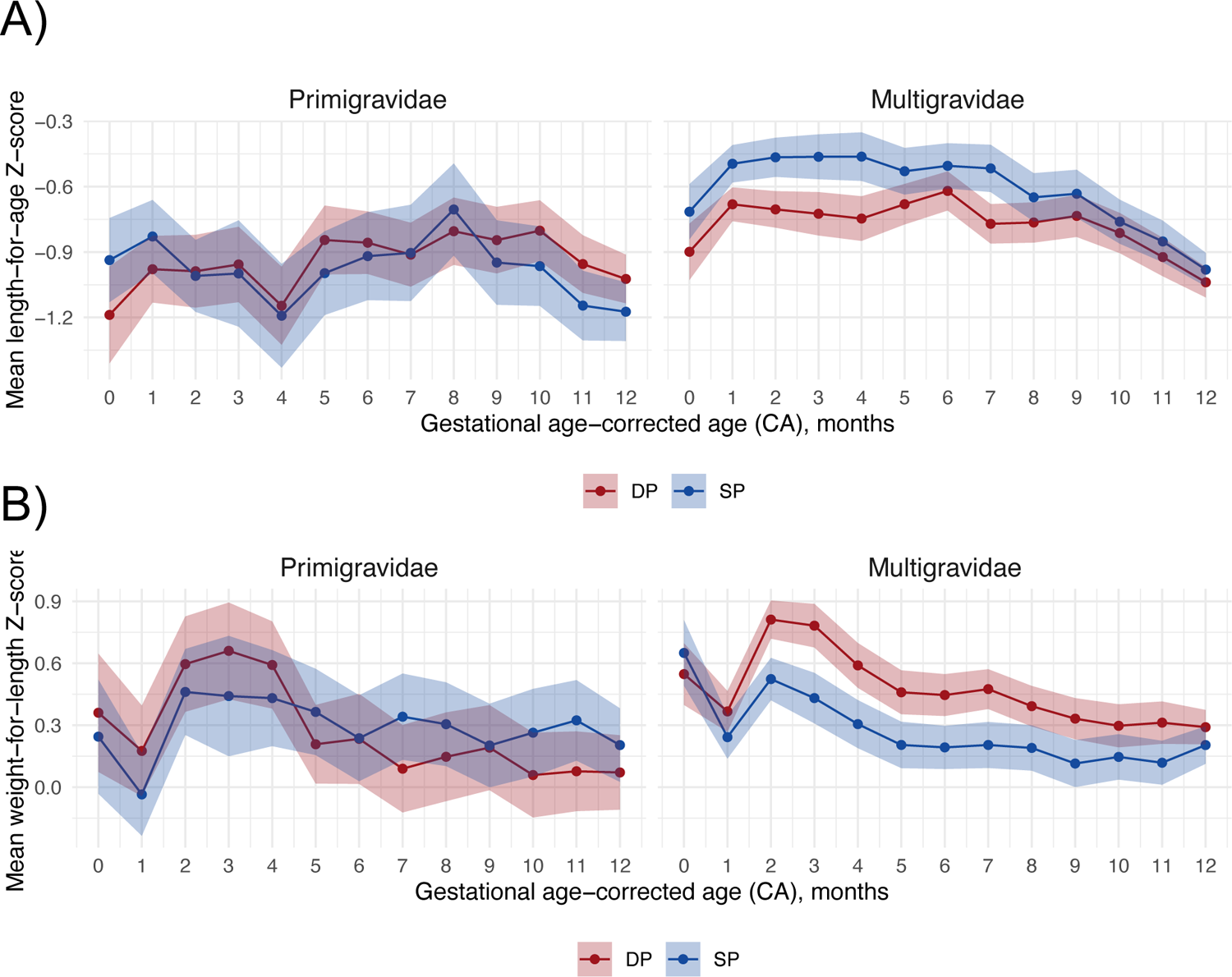
Total effect of IPTp DP vs. SP on mean child growth Z-scores corrected for gestational age by child age and gravidity Includes 630 children measured from birth to 12 months. Excludes children with negative ages after gestational age correction.

**Figure S4.**
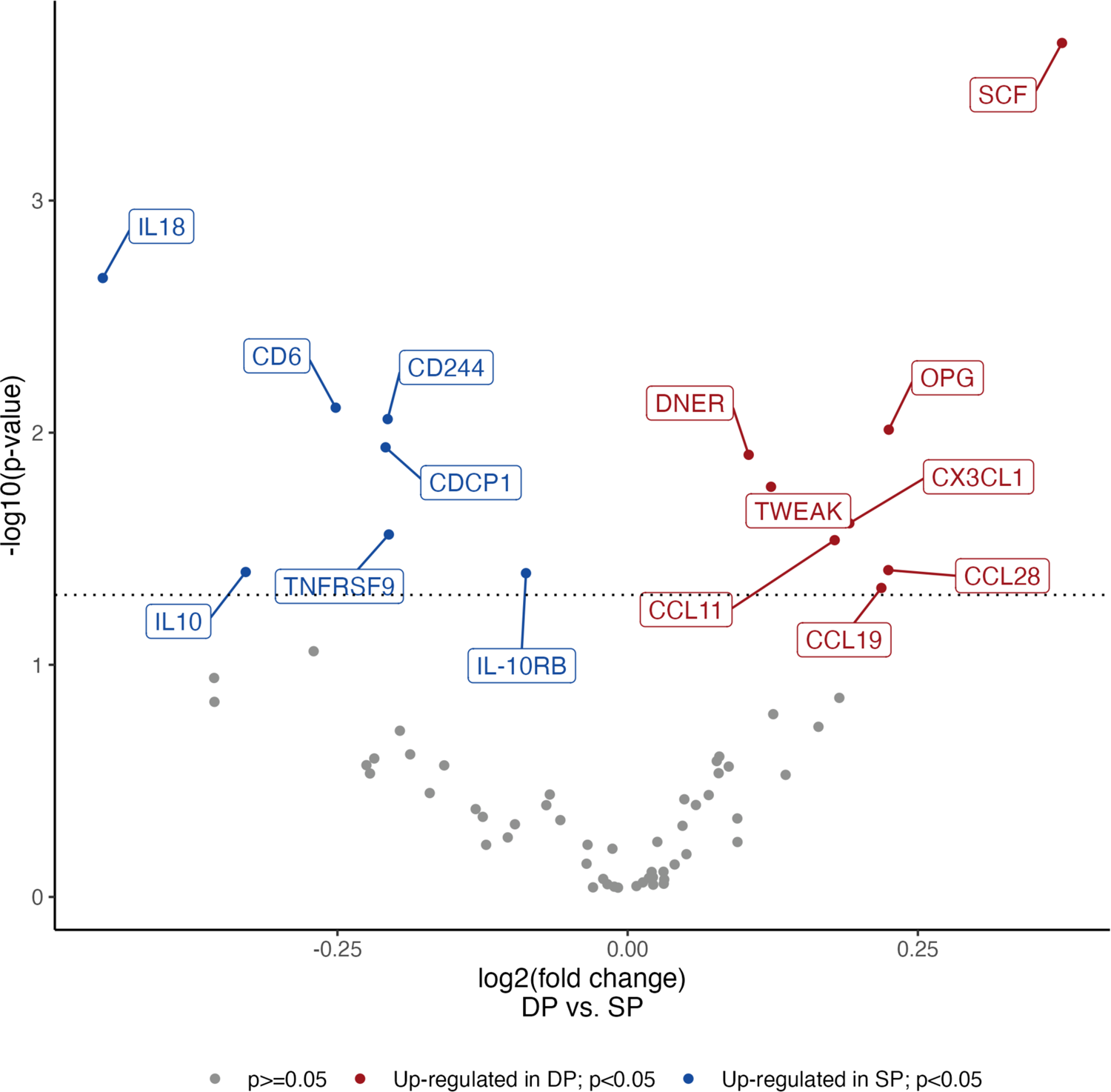
Volcano plot of maternal inflammation-related proteins at delivery by IPTp-DP vs. IPTp-SP among all gravidae Red points represent proteins upregulated in DP with p<0.05, blue points represent proteins upregulated in SP with p<0.05, and gray points represent non-differentially expressed proteins. Color coding in the plot is based on p-values that were not corrected for multiple testing. After the Benjamini-Hochberg correction (false discovery rate p-value < 0.05), only SCF had a p-value < 0.05. Among all gravidae, proteins upregulated in SP were IL18, CD6, CD244, CDCP1, TNFRSF9, IL10, and IL-10RB; proteins upregulated in DP were SCF, OPG, DNER, TWEAK, CX3CL1, CCL11, CCL28, and CCL19. Note that among primigravidae (sample size = 51), proteins CD5 and ADA were upregulated in SP, which were not shown on this volcano plot.

**Figure S5.**
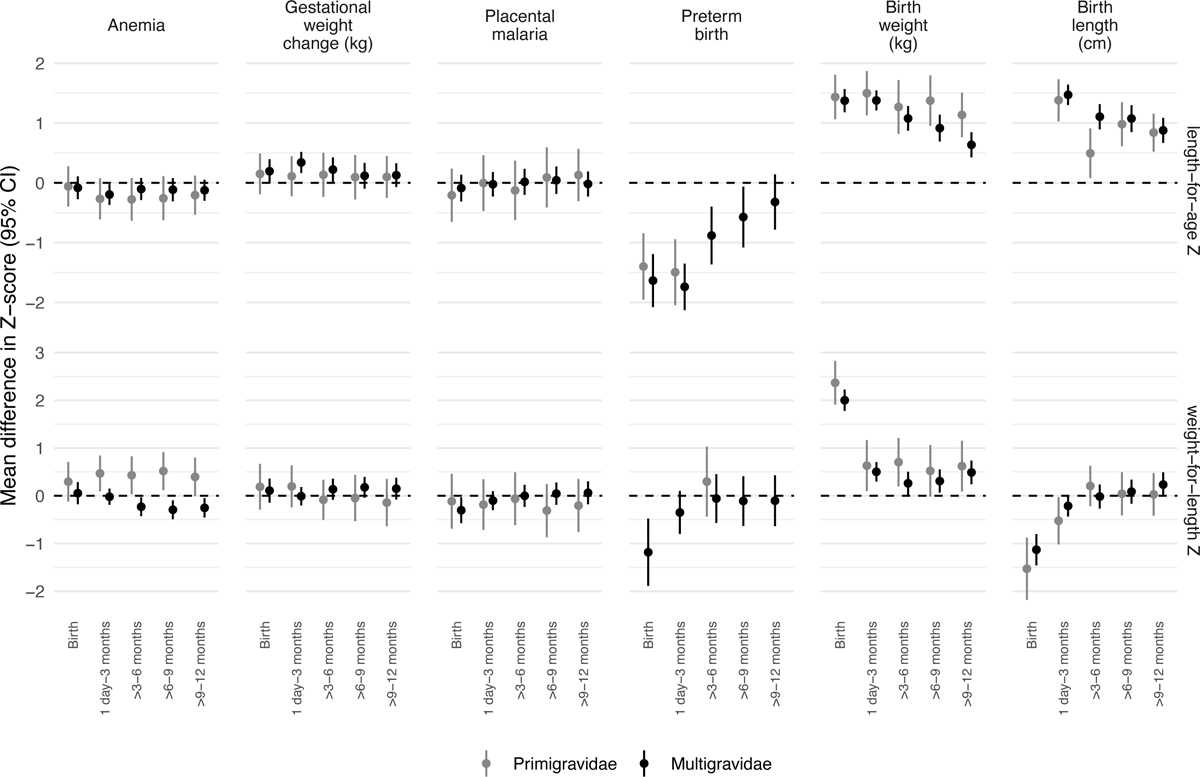
Associations between potential non-inflammation-related mediators and mean child growth Z-scores. Mean differences in Z-scores between each mediator and mean Z-scores adjusted by infant sex, maternal age, maternal baseline parasitemia, gestational age at enrollment, gravidity, maternal education, and household wealth. Note: Gestational weight change and birth length values have been rescaled (multiplied by a factor of 5) to enhance clarity and visualization on the plot. This adjustment aligns their y-axis range with the other mediators for a more visually coherent presentation.

**Figure S6.**
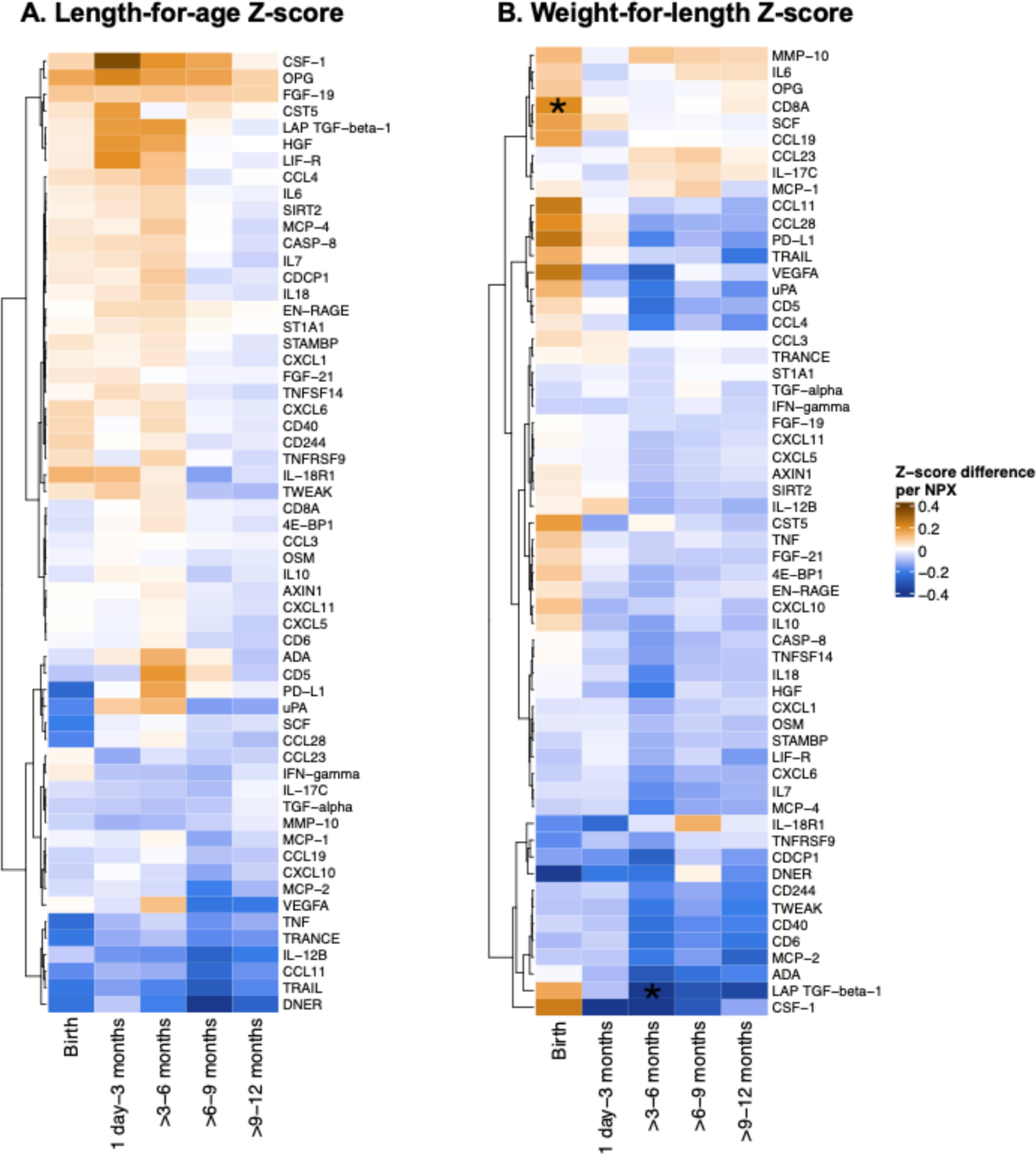
Associations between maternal inflammation-related proteins at delivery and mean child growth Z-scores Mean Z-score differences per inflammation-related protein NPX were estimated with a model adjusted for infant sex, maternal age, gravidity, maternal baseline parasitemia, gestational age at enrollment, maternal education, household wealth. *Statistically significant among all gravidae after Benjamini-Hochberg correction (false discovery rate p-value < 0.05).

**Figure S7.**
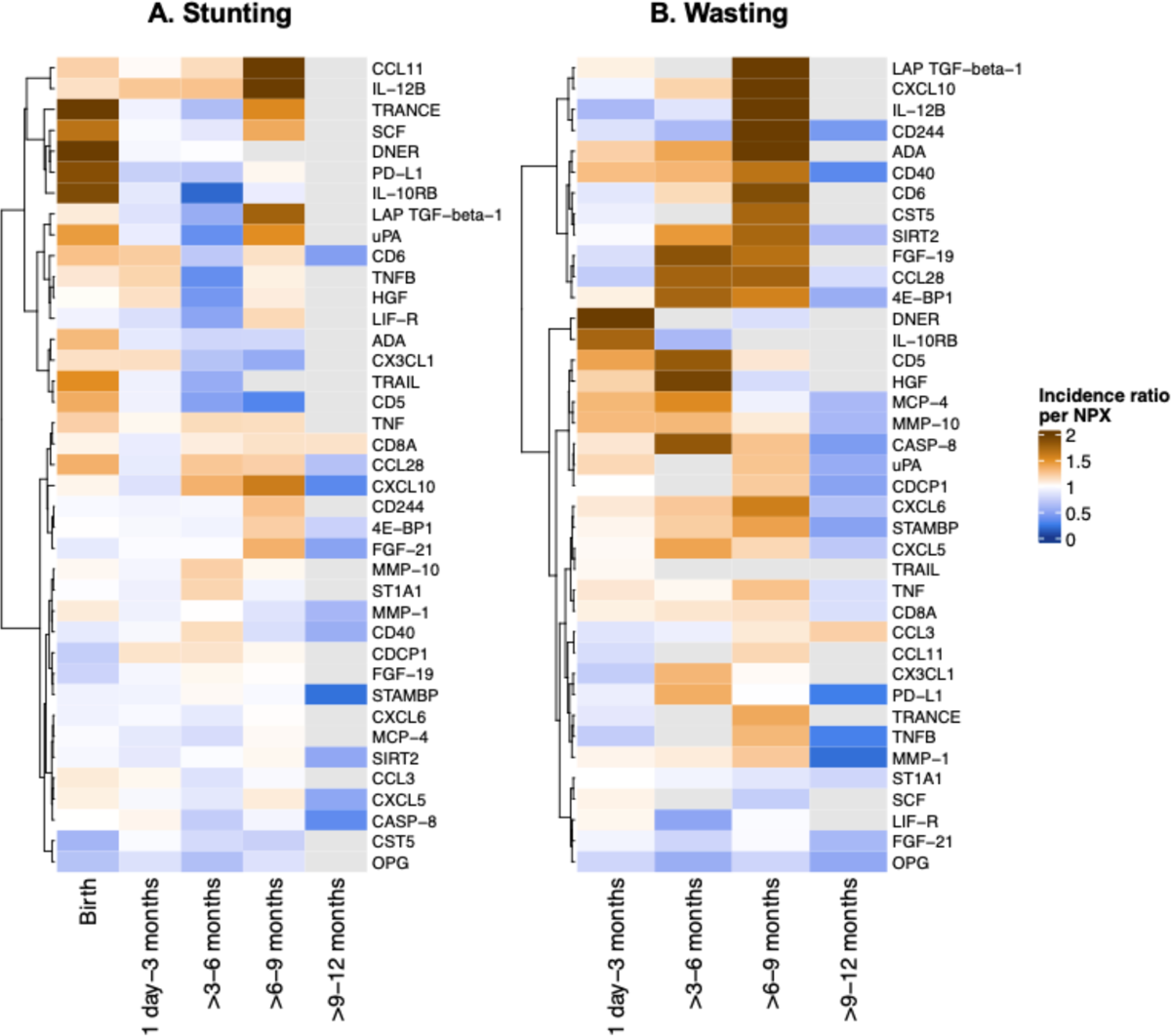
Associations between potential inflammation-related protein mediators and stunting and wasting Incidence ratios per inflammation-related protein NPX were estimated with a model adjusted by infant sex, primigravida, maternal age, maternal baseline parasitemia, gestational age at enrollment, maternal education, and household wealth. The reference group was SP. Missing point estimates were due to data sparsity and were filled as grey. When there were fewer than 10 incident cases in a certain age group, models were not fit.

**Figure S8.**
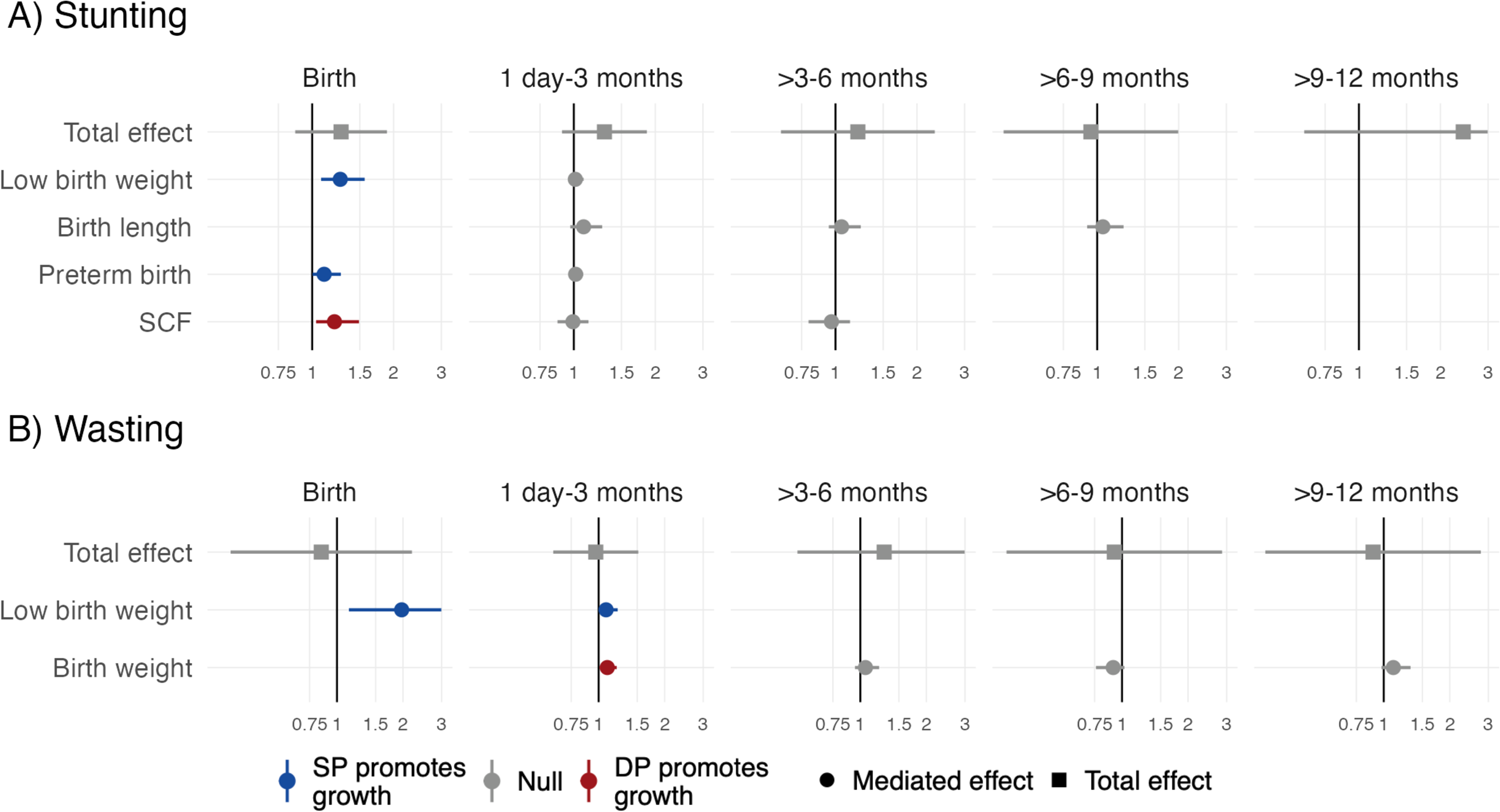
Total effects and mediated effects on incidences of child stunting and wasting The total effects compare incidence of stunting and wasting between IPTp-DP and IPTp-SP using unadjusted models. The mediated effects were adjusted by infant sex, maternal age, maternal baseline parasitemia, gestational age at enrollment, gravidity, maternal education, and household wealth. The reference group was SP. Note: Missing point estimates were due to data sparsity. When there were fewer than 5 incident cases or 5 observed values of a binary mediator in a certain age group, models were not fit.

**Figure S9.**
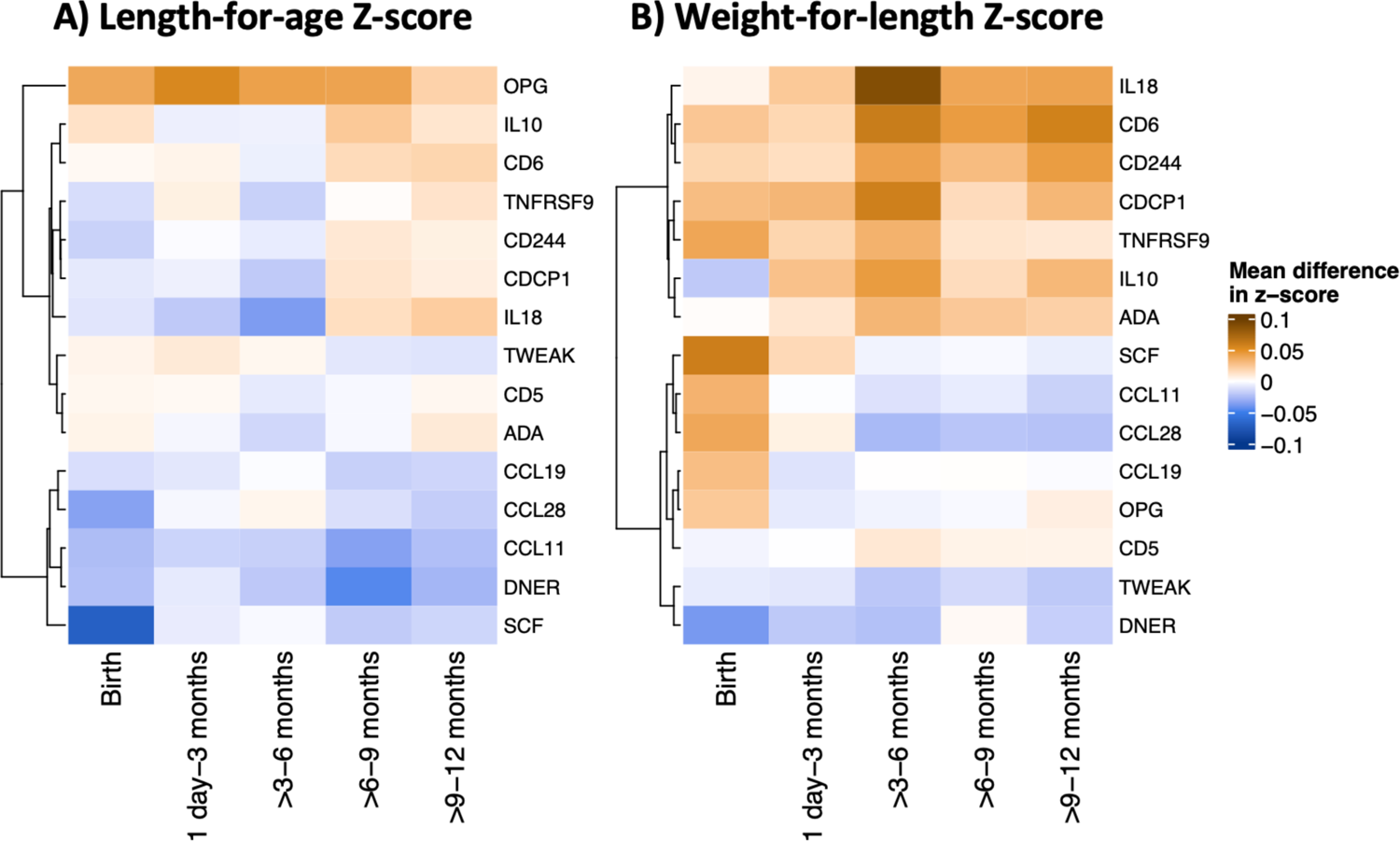
Inflammation-related protein-mediated effects of IPTp-DP vs. IPTp-SP on Z-scores Mediated effects were adjusted by infant sex, primigravida, maternal age, maternal baseline parasitemia, gestational age at enrollment, maternal education, and household wealth. Analysis includes all gravidae. The reference group was SP.

**Figure S10.**
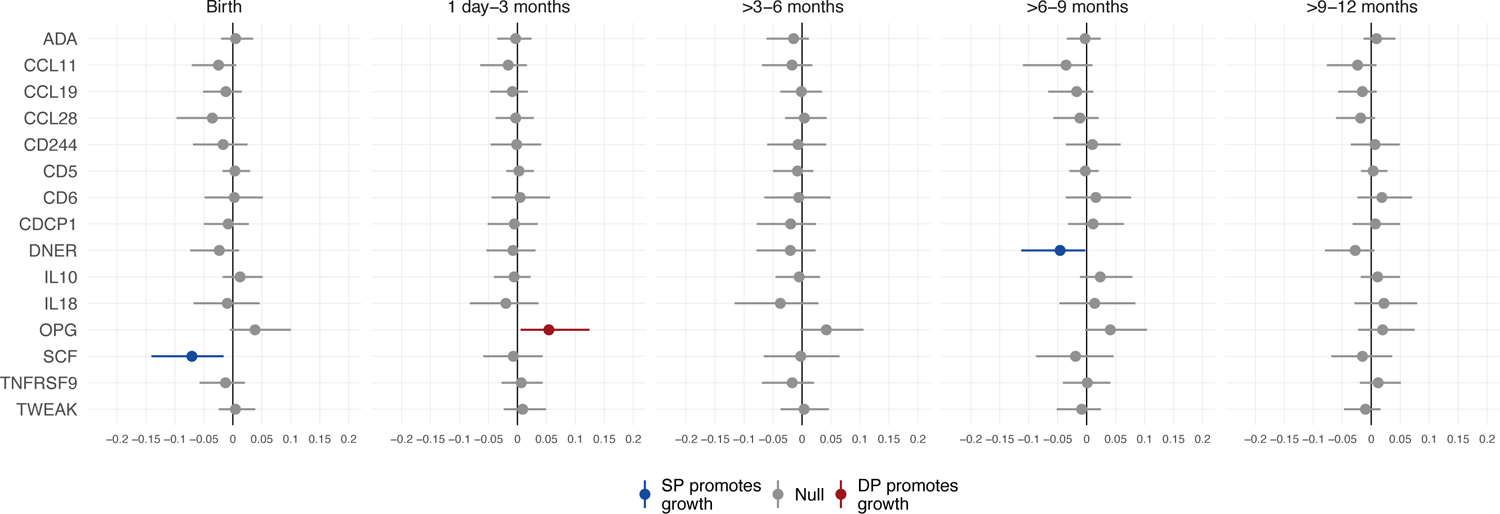
Inflammation-related protein-mediated effects on length-for-age Z Mediated effects were adjusted for infant sex, maternal age, gravidity, maternal baseline parasitemia, gestational age at enrollment, maternal education, and household wealth. Analysis includes all gravidae. The reference group was SP.

**Figure S11.**
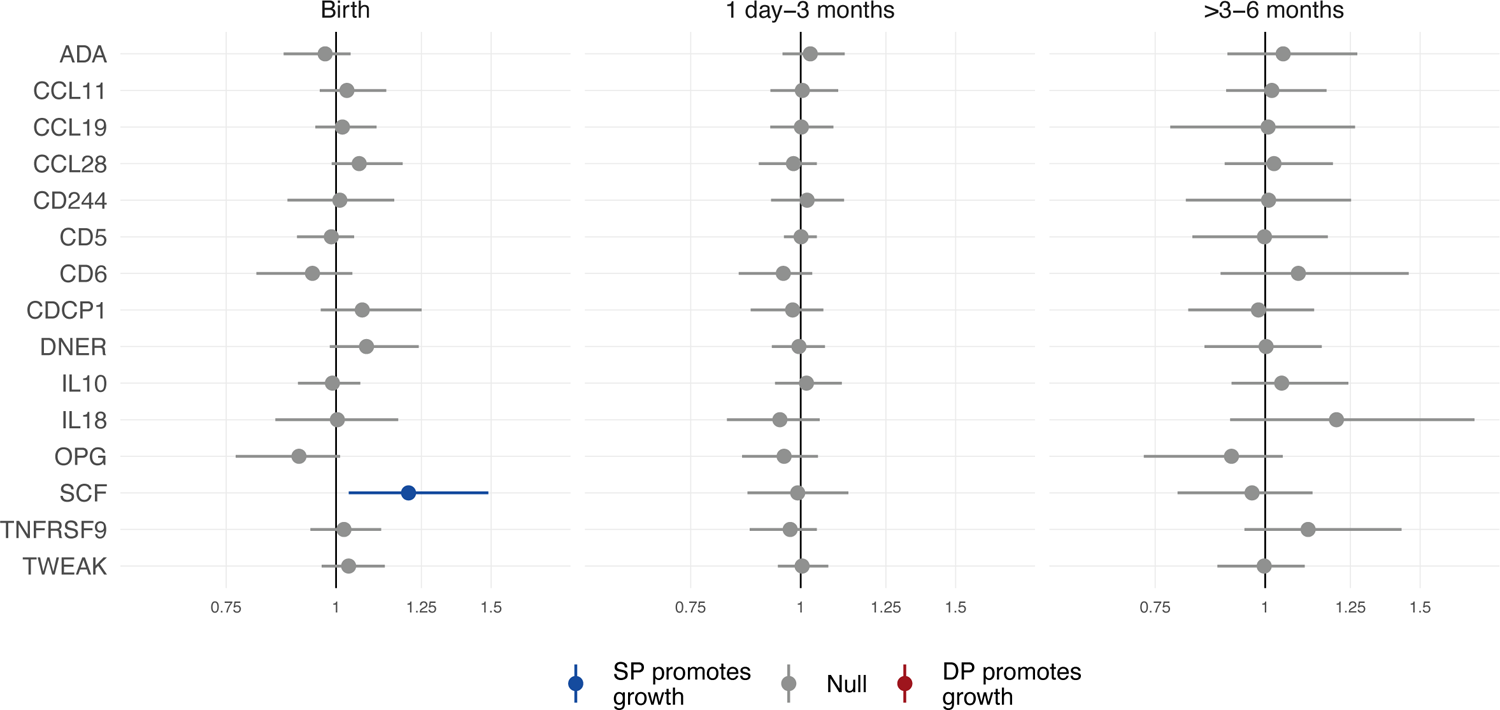
Inflammation-related protein-mediated effects on child stunting Mediated effects were adjusted by infant sex, gravidity, maternal age, maternal baseline parasitemia, gestational age at enrollment, maternal education, and household wealth. Mediation models were only fit from birth through age 6 months due to data sparsity at other ages. Analysis includes all gravidae. The reference group was SP.

**Figure S12.**
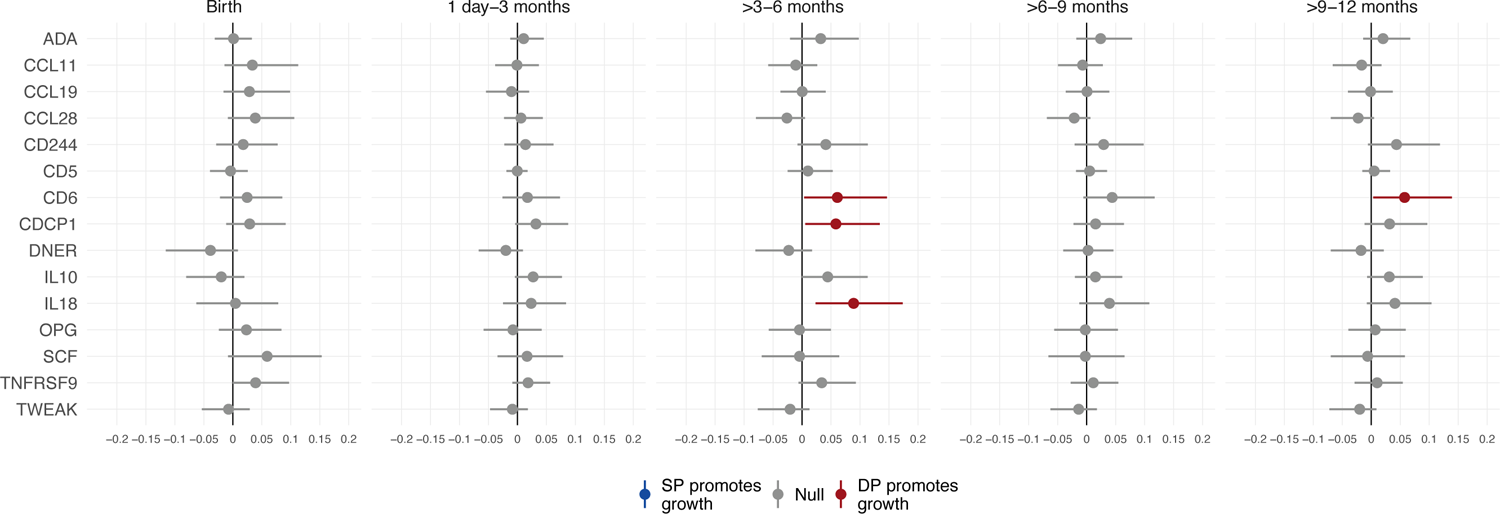
Inflammation-related protein-mediated effects on weight-for-length Z Mediated effects were adjusted by infant sex, gravidity, maternal age, maternal baseline parasitemia, gestational age at enrollment, maternal education, and household wealth. Analysis includes all gravidae. The reference group was SP.

**Figure S13.**
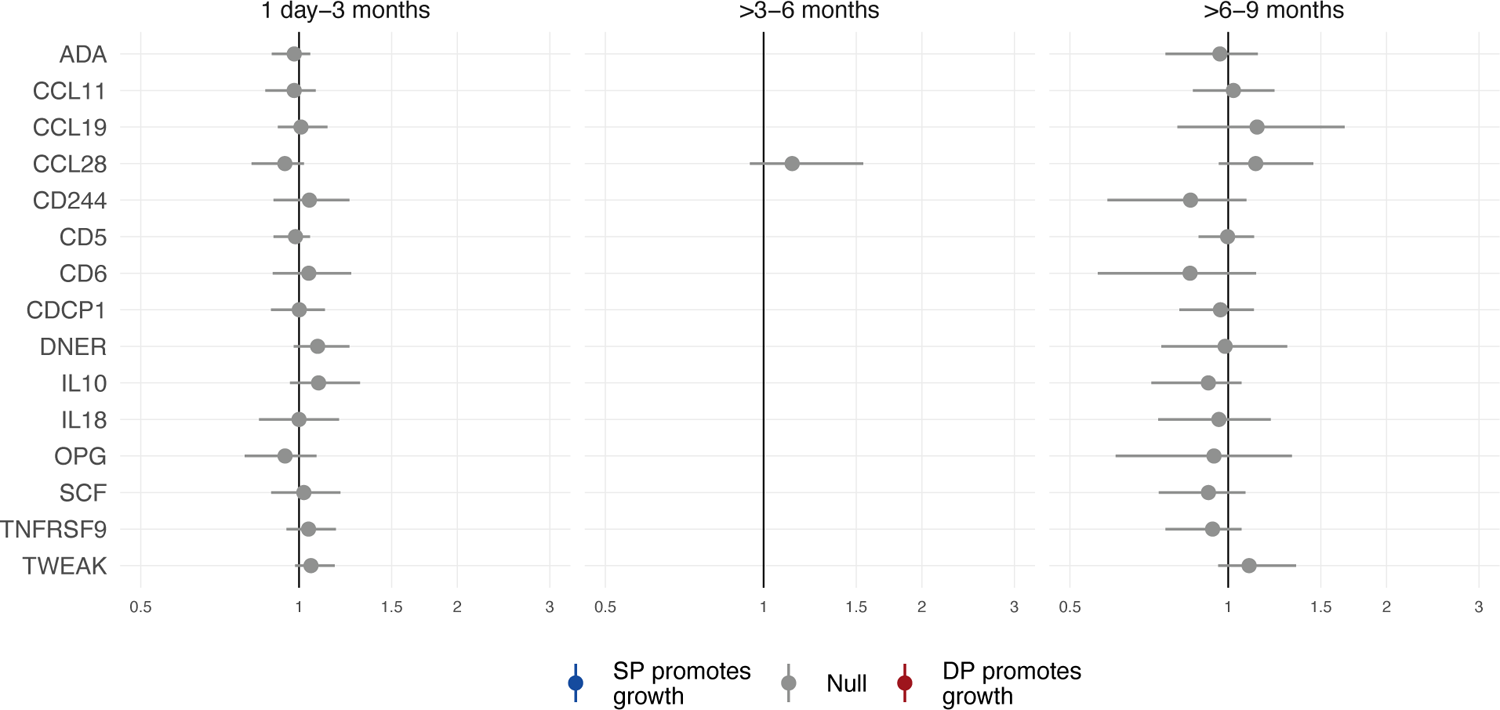
Inflammation-related protein-mediated effects on child wasting. Mediated effects were adjusted by infant sex, gravidity, maternal age, maternal baseline parasitemia, gestational age at enrollment, maternal education, and household wealth. Analysis includes all gravidae. The reference group was SP. Mediation models did not fit at birth or >9-12 months due to data sparsity.

**Table S2.** Total effect of IPTp-DP vs. IPTp-SP on stunting and wasting incidence.

| Age category | N | DP | N | SP | Unadjusted incidence ratio<br>(95% CI) |
| --- | --- | --- | --- | --- | --- |
|  |  | Incidence<br>(95% CI) |  | Incidence<br>(95% CI) |  |
| Stunting |  |  |  |  |  |
| Birth | 324 | 18.2 (14.4, 22.8) | 309 | 14.2 (10.8, 18.6) | 1.28 (0.87, 1.89) |
| 1 day-3 months | 262 | 26.0 (21.0, 31.6) | 260 | 20.0 (15.6, 25.3) | 1.30 (0.90, 1.86) |
| >3-6 months | 182 | 10.4 (6.8, 15.7) | 197 | 8.6 (5.5, 13.4) | 1.21 (0.63, 2.33) |
| >6-9 months | 160 | 8.1 (4.8, 13.4) | 175 | 8.6 (5.3, 13.7) | 0.95 (0.45, 1.99) |
| >9-12 months | 147 | 4.8 (2.3, 9.5) | 153 | 2.0 (0.7, 5.6) | 2.43 (0.63, 9.39) |
| Wasting |  |  |  |  |  |
| Birth | 324 | 2.5 (1.3, 4.8) | 309 | 2.9 (1.5, 5.4) | 0.85 (0.33, 2.20) |
| 1 day-3 months | 312 | 12.5 (9.3, 16.6) | 295 | 12.9 (9.5, 17.2) | 0.97 (0.62, 1.52) |
| >3-6 months | 259 | 4.2 (2.4, 7.4) | 242 | 3.3 (1.7, 6.4) | 1.28 (0.52, 3.19) |
| >6-9 months | 244 | 2.5 (1.1, 5.3) | 225 | 2.7 (1.2, 5.7) | 0.92 (0.30, 2.86) |
| >9-12 months | 236 | 2.5 (1.2, 5.4) | 211 | 2.8 (1.3, 6.1) | 0.89 (0.29, 2.77) |

**Table S3.** Associations between non-inflammation-related potential mediators and stunting and wasting.

|  | Stunting |  | Wasting |  |
| --- | --- | --- | --- | --- |
|  | N | Incidence ratio (95% CI) | N | Incidence ratio (95% CI) |
| Maternal anemia |  |  |  |  |
| Birth | 619 | 1.07 (0.72, 1.58) | 619 | 0.42 (0.14, 1.21) |
| 1 day-3 months | 510 | 1.38 (0.95, 2.00) | 593 | 0.93 (0.59, 1.47) |
| >3-6 months | 374 | 0.83 (0.41, 1.65) | 489 | 1.48 (0.59, 3.70) |
| >6-9 months | 331 | 0.97 (0.45, 2.11) | 459 | 1.51 (0.47, 4.91) |
| >9-12 months | 296 | 0.14 (0.02, 1.11) | 437 | 2.58 (0.74, 9.03) |
| Gestational weight change (kg) |  |  |  |  |
| Birth | 575 | 0.94 (0.85, 1.03) | 575 | 1.05 (0.84, 1.32) |
| 1 day-3 months | 486 | 0.97 (0.89, 1.05) | 552 | 0.93 (0.84, 1.04) |
| >3-6 months | 361 | 0.94 (0.81, 1.09) | 467 | 0.99 (0.80, 1.23) |
| >6-9 months | 321 | 1.06 (0.88, 1.26) | 440 | 0.92 (0.71, 1.19) |
| >9-12 months | 287 | 0.93 (0.71, 1.21) | 419 | 1.01 (0.78, 1.33) |
| Placental malaria |  |  |  |  |
| Birth | 633 | 1.38 (0.85, 2.25) | 633 | 1.14 (0.35, 3.7) |
| 1 day-3 months | 522 | 0.94 (0.60, 1.47) | 607 | 0.98 (0.56, 1.73) |
| >3-6 months | 379 | 1.3 (0.59, 2.86) | 501 | 1.12 (0.35, 3.55) |
| >6-9 months | 335 | 1.21 (0.51, 2.88) | 469 | 0.81 (0.20, 3.38) |
| >9-12 months | 300 | 0.22 (0.02, 1.99) | 447 | 1.01 (0.23, 4.39) |
| Pre-term birth |  |  |  |  |
| Birth | 633 | 3.87 (2.29, 6.52) | 633 | 1.01 (0.13, 7.83) |
| 1 day-3 months | 522 | 2.20 (1.06, 4.56) | 607 | 3.21 (1.71, 6.02) |
| >3-6 months |  | -- |  | -- |
| >6-9 months |  | -- | 469 | 2.85 (0.35, 23.39) |
| >9-12 months |  | -- |  | -- |
| Birth length (cm) |  |  |  |  |
| Birth | 628 | 0.75 (0.72, 0.79) | 628 | 1.33 (1.16, 1.53) |
| 1 day-3 months | 520 | 0.86 (0.81, 0.91) | 605 | 0.98 (0.88, 1.08) |
| >3-6 months | 378 | 0.79 (0.67, 0.92) | 500 | 0.91 (0.74, 1.12) |
| >6-9 months | 334 | 0.79 (0.66, 0.93) | 468 | 1.21 (0.87, 1.68) |
| >9-12 months | 300 | 1.22 (0.87, 1.71) | 446 | 0.9 (0.72, 1.13) |
| Birth weight (kg) |  |  |  |  |
| Birth | 633 | 0.17 (0.12, 0.26) | 633 | 0.15 (0.07, 0.36) |
| 1 day-3 months | 522 | 0.33 (0.22, 0.51) | 607 | 0.35 (0.21, 0.57) |
| >3-6 months | 379 | 0.38 (0.15, 0.97) | 501 | 0.48 (0.13, 1.79) |
| >6-9 months | 335 | 0.27 (0.09, 0.77) | 469 | 3.30 (0.65, 16.68) |
| >9-12 months | 300 | 1.54 (0.26, 9.11) | 447 | 0.24 (0.04, 1.38) |

Incidence ratios for each potential mediator were adjusted by infant sex, primigravida, maternal age, maternal baseline parasitemia, gestational age at enrollment, maternal education, and household wealth.
Note: missing values were due to data sparsity.

**Table S4.** Mediated effects of IPTp-DP vs. IPTp-SP on Z-scores.

|  | Length-for-age Z |  | Weight-for-length Z |  |
| --- | --- | --- | --- | --- |
|  | N | Mean difference in Z (95% CI) | N | Mean difference in Z (95% CI) |
| Maternal anemia |  |  |  |  |
| Birth | 608 | 0.0037 (-0.0077, 0.0225) | 582 | -0.0028 (-0.0229, 0.0103) |
| 1 day-3 months | 593 | 0.0084 (-0.0116, 0.0357) | 581 | -0.0025 (-0.0187, 0.0082) |
| >3-6 months | 575 | 0.0043 (-0.0099, 0.0244) | 574 | 0.0030 (-0.0091, 0.0219) |
| >6-9 months | 559 | 0.0049 (-0.0099, 0.0256) | 558 | 0.0045 (-0.0095, 0.0264) |
| >9-12 months | 548 | 0.0049 (-0.0109, 0.0245) | 548 | 0.0039 (-0.0106, 0.0239) |
| Gestational weight change (kg) |  |  |  |  |
| Birth | 567 | -0.0007 (-0.0182, 0.0144) | 555 | -0.0007 (-0.0183, 0.013) |
| 1 day-3 months | 552 | -0.0036 (-0.0305, 0.0206) | 545 | -0.0002 (-0.0101, 0.0085) |
| >3-6 months | 541 | -0.002 (-0.0264, 0.0186) | 538 | -0.0006 (-0.0139, 0.0092) |
| >6-9 months | 526 | -0.0005 (-0.018, 0.0138) | 525 | -0.0006 (-0.0177, 0.0137) |
| >9-12 months | 515 | 0.0004 (-0.0161, 0.0157) | 515 | 0.0002 (-0.0132, 0.0112) |
| Placental malaria |  |  |  |  |
| Birth | 622 | 0.0358 (-0.0318, 0.1055) | 596 | 0.0903 (-0.002, 0.183) |
| 1 day-3 months | 607 | 0.0049 (-0.0625, 0.0691) | 594 | 0.0414 (-0.0285, 0.1114) |
| >3-6 months | 587 | 0.0003 (-0.075, 0.0708) | 586 | 0.0057 (-0.0706, 0.0805) |
| >6-9 months | 569 | -0.0152 (-0.0897, 0.0545) | 568 | 0.0024 (-0.0727, 0.0782) |
| >9-12 months | 558 | -0.0059 (-0.0745, 0.0588) | 558 | -0.0040 (-0.0778, 0.0691) |
| Pre-term birth |  |  |  |  |
| Birth | 622 | 0.0098 (-0.0645, 0.0909) | 596 | 0.0087 (-0.0252, 0.0511) |
| 1 day-3 months | 607 | 0.0097 (-0.0695, 0.0916) | 594 | 0.0005 (-0.0143, 0.0161) |
| >3-6 months | 587 | 0.0134 (-0.024, 0.0656) | 586 | -0.0013 (-0.0253, 0.0206) |
| >6-9 months | 569 | 0.0112 (-0.0217, 0.0569) | 568 | -0.0017 (-0.0229, 0.0147) |
| >9-12 months | 558 | 0.0087 (-0.0159, 0.0433) | 558 | -0.0033 (-0.0245, 0.0109) |
| Birth length (cm) |  |  |  |  |
| Birth | 622 | -0.2522 (-0.4108, -0.0899) | 596 | 0.0883 (0.0223, 0.1575) |
| 1 day-3 months | 605 | -0.1235 (-0.2265, -0.0319) | 592 | 0.0231 (0.0026, 0.0551) |
| >3-6 months | 585 | -0.0815 (-0.1562, -0.0184) | 584 | -0.0035 (-0.026, 0.017) |
| >6-9 months | 567 | -0.0801 (-0.1586, -0.0099) | 566 | -0.0050 (-0.0254, 0.0123) |
| >9-12 months | 557 | -0.062 (-0.1269, -0.0036) | 557 | -0.0125 (-0.0384, 0.0027) |
| Birth weight (kg) |  |  |  |  |
| Birth | 622 | -0.1115 (-0.2032, -0.023) | 596 | -0.149 (-0.2817, -0.0167) |
| 1 day-3 months | 607 | -0.1119 (-0.2058, -0.0207) | 594 | -0.038 (-0.0786, -0.0045) |
| >3-6 months | 587 | -0.0695 (-0.1444, 0.0009) | 586 | -0.0189 (-0.0499, 0.0037) |
| >6-9 months | 569 | -0.0623 (-0.1333, 0.0031) | 568 | -0.0222 (-0.0541, 0.0005) |
| >9-12 months | 558 | -0.0421 (-0.0943, 0.0059) | 558 | -0.0293 (-0.0707, 0.0036) |
| CD6 |  |  |  |  |
| Birth | 255 | 0.0026 (-0.0485, 0.0519) | 245 | 0.0247 (-0.0221, 0.0853) |
| 1 day-3 months | 251 | 0.0047 (-0.0447, 0.0561) | 247 | 0.0173 (-0.0257, 0.0734) |
| >3-6 months | 255 | -0.0056 (-0.0651, 0.049) | 255 | 0.0609 (0.0037, 0.1464) |
| >6-9 months | 255 | 0.0158 (-0.0361, 0.0765) | 254 | 0.0440 (-0.0056, 0.1172) |
| >9-12 months | 255 | 0.0183 (-0.0237, 0.0704) | 255 | 0.0575 (0.0032, 0.1392) |
| CDCP1 |  |  |  |  |
| Birth | 255 | -0.008 (-0.0500, 0.0274) | 245 | 0.0291 (-0.0115, 0.0911) |
| 1 day-3 months | 251 | -0.0053 (-0.0512, 0.0351) | 247 | 0.0319 (-0.0040, 0.0874) |
| >3-6 months | 255 | -0.0196 (-0.0779, 0.0243) | 255 | 0.0584 (0.0056, 0.1341) |
| >6-9 months | 255 | 0.0111 (-0.032, 0.0643) | 254 | 0.0154 (-0.0227, 0.0643) |
| >9-12 months | 255 | 0.0071 (-0.0321, 0.0496) | 255 | 0.0316 (-0.0115, 0.0965) |
| DNER |  |  |  |  |
| Birth | 255 | -0.0233 (-0.0734, 0.0107) | 245 | -0.0386 (-0.1157, 0.0088) |
| 1 day-3 months | 251 | -0.0077 (-0.0532, 0.0312) | 247 | -0.0200 (-0.0668, 0.0095) |
| >3-6 months | 255 | -0.0202 (-0.0785, 0.0234) | 255 | -0.023 (-0.0805, 0.0173) |
| >6-9 months | 255 | -0.0458 (-0.1128, -0.0026) | 254 | 0.0024 (-0.0404, 0.0461) |
| >9-12 months | 255 | -0.0277 (-0.0798, 0.0054) | 255 | -0.0177 (-0.0699, 0.0215) |
| IL18 |  |  |  |  |
| Birth | 255 | -0.0095 (-0.0678, 0.0465) | 245 | 0.0046 (-0.0630, 0.0784) |
| 1 day-3 months | 251 | -0.0199 (-0.0822, 0.0362) | 247 | 0.0237 (-0.0248, 0.0838) |
| >3-6 months | 255 | -0.0374 (-0.1161, 0.0281) | 255 | 0.0890 (0.0234, 0.1738) |
| >6-9 months | 255 | 0.0139 (-0.0470, 0.0840) | 254 | 0.0393 (-0.0126, 0.1082) |
| >9-12 months | 255 | 0.0219 (-0.0291, 0.0792) | 255 | 0.0408 (-0.0076, 0.1038) |
| OPG |  |  |  |  |
| Birth | 255 | 0.0384 (-0.0049, 0.1003) | 245 | 0.0235 (-0.0239, 0.0839) |
| 1 day-3 months | 251 | 0.0543 (0.0057, 0.1243) | 247 | -0.0077 (-0.0583, 0.0418) |
| >3-6 months | 255 | 0.0419 (-0.0025, 0.106) | 255 | -0.0044 (-0.0573, 0.0497) |
| >6-9 months | 255 | 0.0408 (-0.0026, 0.1041) | 254 | -0.0023 (-0.0559, 0.0538) |
| >9-12 months | 255 | 0.0196 (-0.0228, 0.0747) | 255 | 0.0068 (-0.0395, 0.0594) |
| SCF |  |  |  |  |
| Birth | 255 | -0.0706 (-0.1404, -0.0161) | 245 | 0.0592 (-0.0082, 0.1535) |
| 1 day-3 months | 251 | -0.0071 (-0.0587, 0.0435) | 247 | 0.0166 (-0.0342, 0.0787) |
| >3-6 months | 255 | -0.0022 (-0.0658, 0.0646) | 255 | -0.0043 (-0.0696, 0.0641) |
| >6-9 months | 255 | -0.0192 (-0.0874, 0.0463) | 254 | -0.0023 (-0.0660, 0.0652) |
| >9-12 months | 255 | -0.0152 (-0.0689, 0.0361) | 255 | -0.0063 (-0.0699, 0.0581) |

Mediated effects were adjusted for infant sex, primigravida, maternal age, maternal baseline parasitemia, gestational age at enrollment, maternal education, and household wealth. Analysis includes all gravidae. The reference group was SP.

## Notes

### Competing Interest Statement

The authors have declared no competing interest.

